# America Addresses Two Epidemics – Cannabis and Coronavirus and their Interactions: An Ecological Geospatial Study

**DOI:** 10.1101/2020.04.17.20069021

**Authors:** Albert Stuart Reece, Gary Kenneth Hulse

## Abstract

**Importance:** Covid-19 infection has major international health and economic impacts and risk factors for infection are not completely understood. Cannabis smoking is linked with poor respiratory health, immunosuppression and multiple contaminants. Potential synergism between the two epidemics would represent a major public health convergence. Cigarettes were implicated with disease severity in Wuhan, China.

**Objective:** Is cannabis use epidemiologically associated with coronavirus incidence rate (CVIR)?

**Design:** Cross-sectional state-based multivariable study.

**Setting:** USA.

**Primary and Secondary Outcome Measures:** CVIR. Multivariable-adjusted geospatially-weighted regression models. As the American cannabis epidemic is characterized by a recent doubling of daily cannabis use it was considered important to characterize the contribution of high intensity use.

**Results:** Significant associations of daily cannabis use quintile with CVIR were identified with the highest quintile having a prevalence ratio 5.11 (95%C.I. 4.90-5.33), an attributable fraction in the exposed (AFE) 80.45% (79.61-81.25%) and an attributable fraction in the population of 77.80% (76.88-78.68%) with Chi-squared-for-trend (14,782, df=4) significant at P<10^−500^. Similarly when cannabis legalization was considered decriminalization was associated with an elevated CVIR prevalence ratio 4.51 (95%C.I. 4.45-4.58), AFE 77.84% (77.50-78.17%) and Chi-squared-for-trend (56,679, df=2) significant at P<10^−500^. Monthly and daily use were linked with CVIR in bivariate geospatial regression models (P=0.0027, P=0.0059). In multivariable additive models number of flight origins and population density were significant. In interactive geospatial models adjusted for international travel, ethnicity, income, population, population density and drug use, terms including last month cannabis were significant from P=7.3×10^−15^, daily cannabis use from P=7.3×10^−11^ and last month cannabis was independently associated (P=0.0365).

**Conclusions and Relevance:** Data indicate CVIR demonstrates significant trends across cannabis use intensity quintiles and with relaxed cannabis legislation. Recent cannabis use is independently predictive of CVIR in bivariate and multivariable adjusted models and intensity of use is interactively significant. Cannabis thus joins tobacco as a SARS2-CoV-2 risk factor.

**Article Summary:** Strengths and Limitations of this Study

- Population level was used for the large datasets employed relating to international travel, Covid-19 rates and drug exposure.
- Nationally representative datasets were employed for drug use and exposure
- A Broad range of covariates was considered including socioeconomic, demographic, drug use, Covid-19 incidence and international travel.
- Advanced geospatial modelling techniques were used to analyze data.
- Higher resolution geospatial data was not available to this study.

**Note:** The following files were submitted by the author for peer review, but cannot be converted to PDF.You must view these files (e.g. movies) online.

**Key Points:** *Question:* Since cannabis is immunosuppressive and is frequently variously contaminated, is its use associated epidemiologically with coronavirus infection rates?

*Findings:* Geospatial analytical techniques were used to combine coronavirus incidence, drug and cannabinoid use, population, ethnicity, international flight and income data. Cannabis use and daily cannabis use were associated with coronavirus incidence on both bivariate regression and after multivariable spatial regression with high levels of statistical significance. Cannabis use quintiles and cannabis legal status were also highly significant.

*Meaning:* Significant geospatial statistical associations were shown between cannabis use and coronavirus infection rates consistent with immunomodulatory mechanistic reports and environmental exposure concerns.

**BMJ:** *I, the Submitting Author has the right to grant and does grant on behalf of all authors of the Work (as defined in the below author licence), an exclusive licence and/or a non-exclusive licence for contributions from authors who are: i) UK Crown employees; ii) where BMJ has agreed a CC-BY licence shall apply, and/or iii) in accordance with the terms applicable for US Federal Government officers or employees acting as part of their official duties; on a worldwide, perpetual, irrevocable, royalty-free basis to BMJ Publishing Group Ltd (“BMJ”) its licensees and where the relevant Journal is co-owned by BMJ to the co-owners of the Journal, to publish the Work in this journal and any other BMJ products and to exploit all rights, as set out in our [licence](https://authors.bmj.com/wp-content/uploads/2018/11/BMJ_Journals_Combined_Author_Licence_2018.pdf)*.

*The Submitting Author accepts and understands that any supply made under these terms is made by BMJ to the Submitting Author unless you are acting as an employee on behalf of your employer or a postgraduate student of an affiliated institution which is paying any applicable article publishing charge (“APC”) for Open Access articles. Where the Submitting Author wishes to make the Work available on an Open Access basis (and intends to pay the relevant APC), the terms of reuse of such Open Access shall be governed by a Creative Commons licence – details of these licences and which [Creative Commons](http://creativecommons.org/) licence will apply to this Work are set out in our licence referred to above*.

*Other than as permitted in any relevant BMJ Author’s Self Archiving Policies, I confirm this Work has not been accepted for publication elsewhere, is not being considered for publication elsewhere and does not duplicate material already published. I confirm all authors consent to publication of this Work and authorise the granting of this licence*.

## Introduction

The coronavirus pandemic of January-March 2020 has gathered great attention worldwide and is accelerating globally at the time of writing. With a mortality originally posted by WHO at 3-4% ^1^ rising to over 10% in some nations ^2^, and ventilator shortages reported in Italy ^3^ and USA ^4,5^ there is considerable cause for concern. Importantly whilst senior NIH authorities have since revised mortality estimates in the general population downward to below one percent ^6^ mortality rates in the elderly and patients with chronic disease are likely to remain appreciable ^6-9^. Coronavirus data on March 27^th^ 2020 showed that there had been 94,014 cases and 1,431 deaths attributed to the virus in USA to that time (1.52% mortality) ^10^.

When risk factors for severe infection with coronavirus were recently been studied in Hubei province on China in three tertiary hospitals in Wuhan, tobacco smoking was identified in 27.3% of patients with progressive disease v 3% of non-progressive disease (N= 11 progressors and 67 non-progressors, P=0.018) ^7^.

Importantly the cannabis industry is known to have recently increased its activity significantly in USA following widespread relaxation of regulations pertaining to its use, and a 2018 study indicated that legalization was associated with an increase of more than 1,000,000 cannabis users and 500,000 cannabis-dependent people ^11^. A large literature describes the immunosuppressive properties of several cannabinoids including Δ9-tetrahydrocannabinol (THC), cannabidiol and cannabinol ^12-20^. Cannabis users frequently inhale with deep breaths which are held for long period so that smoke can penetrate deeply into the lung ^12,21^. Moreover cannabis has been shown to be contaminated with foreign chemicals, viruses and fungal spores ^22-25^ so that concern has been expressed that patients can be relatively immunocompromised and at heightened risk of exposure to microorganisms placing them at increased risk of infection ^26^.

Moreover the recent World Drug Report 2019 released from the Office of Drugs and Crime of United Nations emphasized that whilst the US revival of cannabis is including more users, it is primarily about an increase in the number of daily or near daily users with that rate having doubled 2008-2018 ^27^. It is important to consider then that we can expected to see more patients presenting with the effects of high level cannabis use. This important datum suggests that the immunosuppressive effects of cannabis are likely to be magnified in habitual users by the higher potency of modern strains, increased exposure and deep inhalation smoking habits in this context.

There are therefore a number of theoretical reasons for being concerned that cannabis use may exacerbate infectious risks such as that posed by coronavirus as was recently suggested^26^.

The present study is an ecological exploration designed to test the hypothesis that there may be epidemiological evidence for a geospatial association between high rates of cannabis use with increased coronavirus infection rates (CVIR). The study was performed based on USA data as that nation has the best publicly available datasets available which allow formal analysis. The hypothesis was formulated prior to study commencement.

## Methods

Data. U.S. state-based Corona virus data was taken from the worldometer website on March 27^th^ 2020 ^10^. The most recently available data on 49,320 international flights into USA (from October 2018-September 2019) was taken from the Department of Transport ^24^. Drug use by state data was taken from the 2017-2018 Restricted Use Data Analysis System (RDAS) held by the Substance abuse and Mental Health Services Administration (SAMHSA) ^28^. Eight drug codes were employed namely: IRABUPOSPNR for prescription pain reliever abuse in past year, PNRNMYR for Recoded – pain relievers past year misuse, MRJMDAYS for percent using cannabis on all or most days (defined as ≤20 days per month), MJRMON for past month cannabis use, COCYR for past year cocaine use, CIGMON for past month cigarette use, BNGALC for binge alcohol use in past month and AMPHETAPYU for any amphetamine past year use. State recreational cannabis legal status was taken from an internet search ^29^. State population, ethnicity and median household income was derived from the US Census Bureau five year American Community Survey (ACS) for 2018 via the tidycensus package in R. State area is included in the albersusa R package and was used to derive population density.

### Data Sharing Statement

Study data is made available with this paper in the online supplementary material.

### Statistics

Data was processed in RStudio version 1.2.1335 based on R version 3.6.1 on 1^st^ April 2020. Parameters were log transformed depending on the results of the Shapiro test. The packages dplyr, sf, albersusa, spdep, splm were used for data import, manipulation, analysis and drawing of maps and graphs. Non-parametric analysis was performed using the Wilcoxson test. Chi squared test for trend was done in R. Prevalence ratios and associated measures were calculated using the epiR::epi9.2by2 function. Geospatial interstate queen-based (edge and corner) links were derived with spdep::poly2nb and edited manually as indicated. Geospatial analysis was performed in the package splm using spatial panel maximum likelihood (spml) and spatial panel generalized method of moments (spgm) by Millo and Piras ^30^ and spatial panel random effects maximum likelihood (spreml) analysis was performed using splm::spreml ^31^. The spatial error structure used for spml models was that of Kapoor, Kelejian and Prucha (KKP) ^32^. Further details are given in the Tables. spml models were compared using the spatial Hausman test (sphtest) with directionality informed by the Log Likelihood Ratio (logLik). The spatial error structure used in spreml models was the full error structure (spatial error after Kapoor Kelejian and Prucha with serially correlated remainder errors and random effects (sem2srre) without lagging). The appropriateness of this error structure was formally tested by substituting various alternative forms and comparing results including the logLik. P<0.05 was considered significant.

### Patient and Public Involvement Statement

Patients were involved in this research at several points. Patients worldwide are very concerned about the Covid-19 epidemic and the implications for their health, their lifespan, their quality of life, their risk of unemployment and many serious matters related to this. Patients are also concerned about the things they can do to stay healthy. Patients are concerned about possible risk factors for health and well being. For this reason they are interested in the subject of the present investigation. Patients have also been most interested in the results. They are interested in how they can apply this result to their own lives and to that of friends and family who might be close to them.

Hence the research questions and outcome measures were developed and informed by patients priorities, experiences and preferences. Our patients were involved in the design of this study in that they unanimously agreed that such matters should be investigated from extant publicly accessible databases. Patients were not involved in patient recruitment as that was not applicable to a study of this methodology. Hence their time was not consumed with the actual conduct and performance of this research.

Patients have been widely consulted about the best way to disseminate the results of this research. The agreed that publication in reputable professional medical journals is advisable and preferable. They also feel that such efforts should be supported on social media and on mainstream media to the extent that commercial radio personalities might be interested in such subjects for indeed at the time of writing the coronavirus pandemic is receiving very extensive media coverage indeed.

### Ethics

This study was approved by the Human Research Ethics Committee of the University of Western Australia on 31^st^ March 2020 (No. RA/4/20/4724).

## Results

Figure 1 shows the rates of coronavirus infection (A) and death (B) by state across USA as of March 27^th^ 2020.

**Figure 1.**
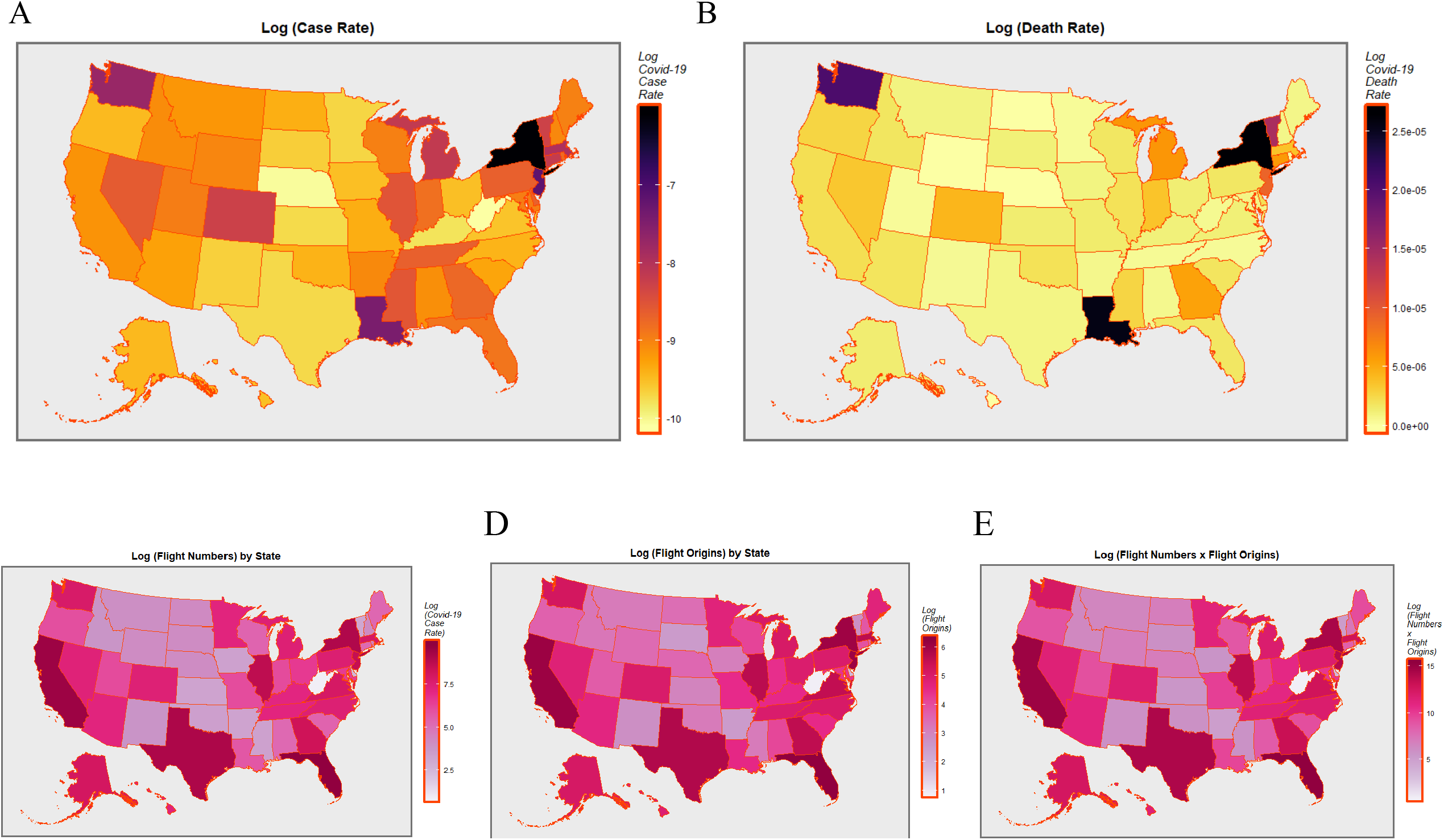
Covid-19 Rates and Flight Data. (A) Choropleth map of log (Case Rate) by US State. (B) Choropleth map of log (Mortality Rate) by US State. (C) Choropleth map of log (Flight Numbers) by US State. (D) Choropleth map of log (Numbers of Flight Origins) by US State. (E) Choropleth map of log (Product of Flight Numbers x Numbers of Flight Origins) by US State.

It is known that air travel is one of the primary vectors of spread of the virus. For this reason it was of interest to quantitate this. The most recent data on international flights to USA from the US Department of Transport was sourced and is also shown map-graphically in Figure 1 showing the number of flights (Figure 1C), numbers of flight origins (Figure 1D), and the product of these two parameters (Figure 1E).

Other state-based socioeconomic data including population, area, population density and median household income were also sourced from the US Census Bureau (USCB) and shown in Supplementary Figure 1. Ethnicity data sourced from USCB is shown in Supplementary Figure 2.

State-based drug use data was sourced from the RDAS maintained by SAMHSA relating to the use of cigarettes, binge alcohol, amphetamines, opioids and cocaine. Two metrics of cannabis use were obtained related to any use the past month (MRJMON) and percent smoking cannabis daily or near daily (≥20 days/month, MRJMDAYS; denoted hereafter “daily cannabis use”). This data is shown map-graphically in Figure 2.

**Figure 2.**
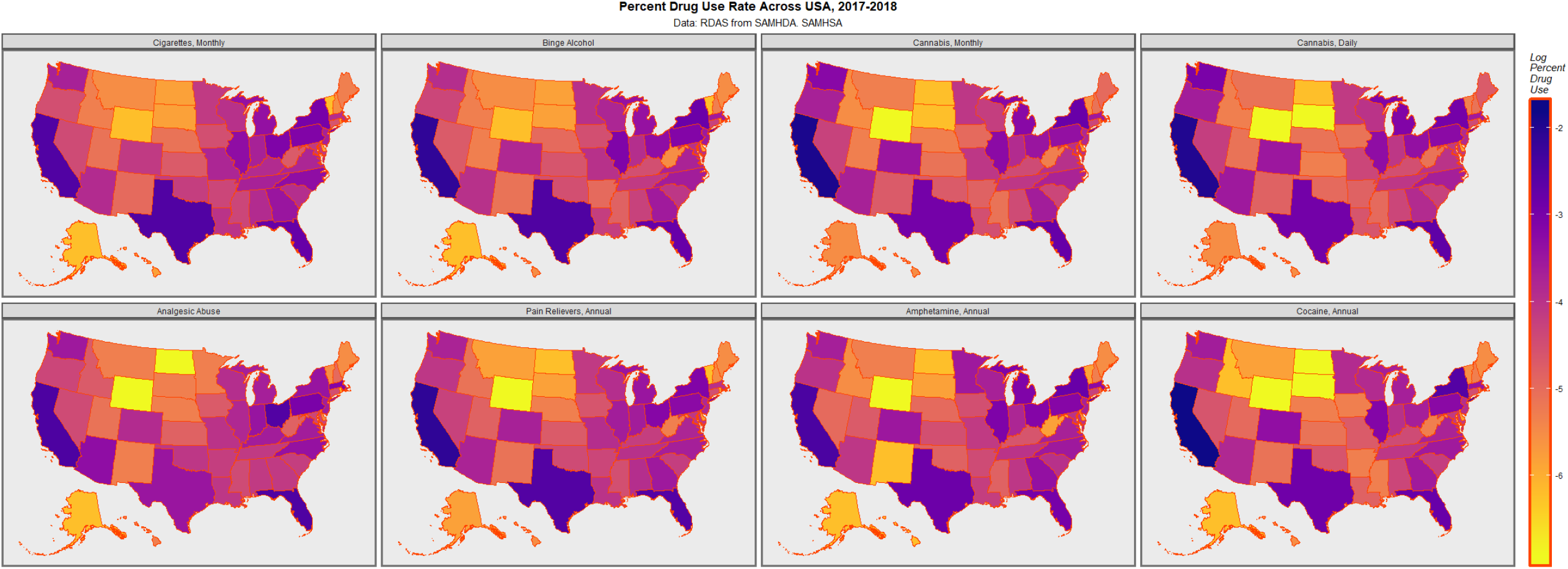
Choropleth maps of log(Drug Use Rates) by US State. Data, Restricted Use Data Analysis System (RDAS) from Substance Abuse and Mental Health Services Data Archive (SAMHDA) from SAMHSA ^28^.

Supplementary Table 1 provides a tabulation of the states by their daily cannabis use quintile and the legal status of cannabis in 2020. Figure 3A shows a boxplot of the CVIR by quintile of daily cannabis use with Quintile 5 being the lowest daily use and Quintile 1 being the highest daily use. Whilst the trend appears to be positively skewed the notches of the boxes overlap indicating lack of statistical difference. Supplementary Table 2 shows the Prevalence ratio (PR, like odds ratio for cross-sectional data), the attributable fraction in the exposed (AFE) and the attributable fraction in the population (AFP) calculated numerically directly from the case numbers. As can be seen the PR’s rise monotonically with Quintile number from 1.22 (95%C.I. 1.14-1.31) to 5.11 (4.90-5.33). The AFE’s rise from 18.15% (12.49-23.44%) to 80.45 (79.61-81.25%) and the AFP’s rise from 6.9% (4.51-9.24%) to 77.80% (76.88-78.68%). These are very significant fractions indeed. (Chi-squared for trend = 14,782, df=4, P<2.2×10^−500^; for comparison χ^2^ =1,478, df=4, P=8.43×10^−319^).

**Figure 3.**
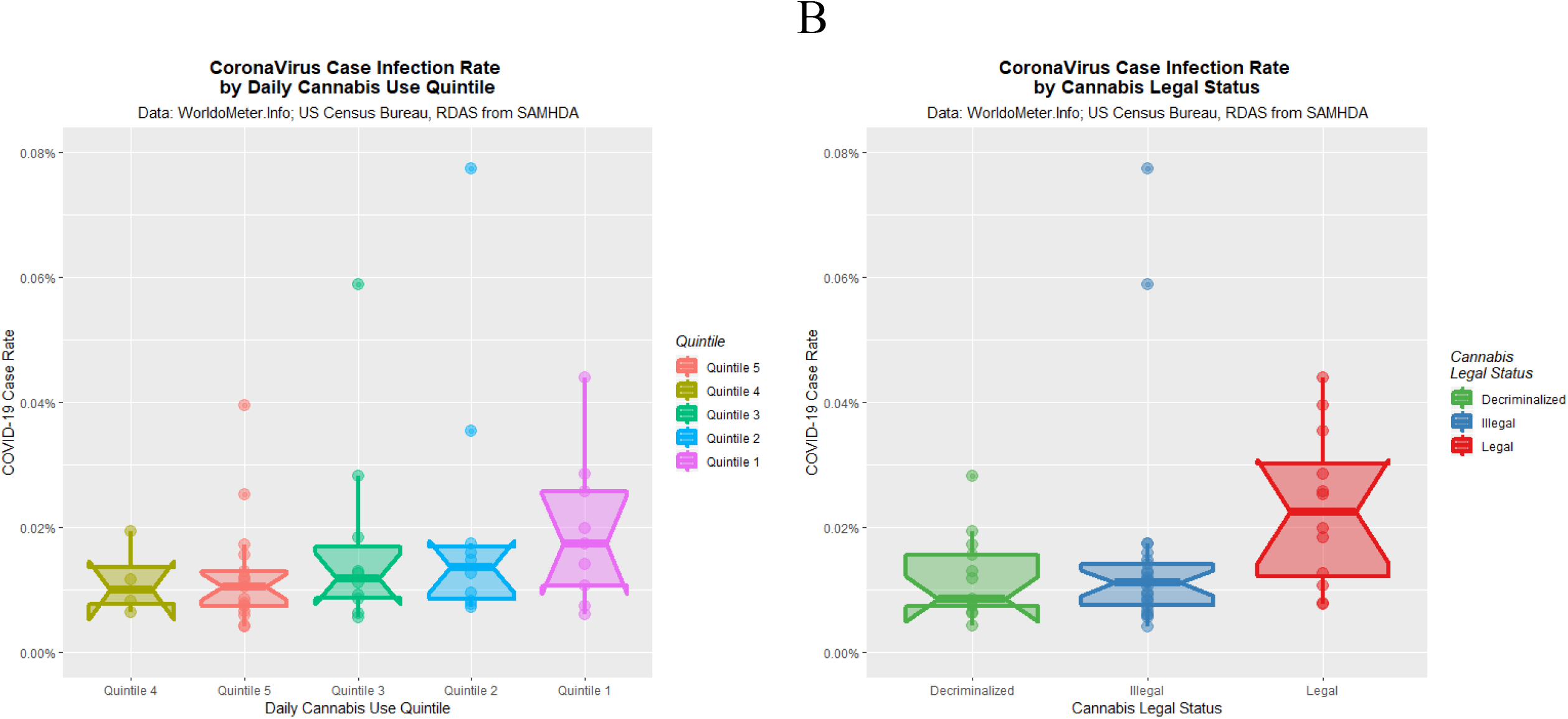
Impact of Cannabis Daily Use Quintiles and Legal Status on Coronavirus Infection Rates. (A) Coronavirus infection rate by daily cannabis use quintiles. (B) Coronavirus infection rate by recreational cannabis use legal status.

Figure 3B shows the coronavirus infection rate as a function of the legal status of recreational cannabis. At this time cannabis is legal in 11 states, illegal in 25 states and decriminalized in 14 states. This Figure is calculated from the CVIR. Again a positive increment is noted with relaxation of cannabis regulations. This time however the notches do not overlap. Supplementary Table 2 shows the case rates calculated from the raw case numbers. Rising PR’s, AFE’s and AFP’s are noted. Cannabis decriminalization is noted in this analysis to be associated with a PR of 4.51 (4.45-4.58), an AFE of 77.84% (77.50-78.17%) and an AFP of 51.22% (50.74-51.70%). (Chi-squared for trend = 56,679, df=2, P<2.2×10^−500^; for comparison χ^2^ =567, df=2, P=7.54×10^−124^). These data look different from those in Figure 3B due to the skewing effect of outliers. When non-parametric analysis was used on these CVIR the illegal-legal difference was significant (W=72, P=0.0239) but the illegal– decriminalized difference was not (W=180, P = 0.8965).

It was therefore of interest to consider these data from a geospatial analytical perspective. Supplementary Figure 3A shows the links derived from the spdep::poly2nb function and how these were edited to allow Alaska to conceptually relate to Washington state and Oregon and Hawaii to California. The final neighbour link network used is shown in Supplementary Figure 3B.

Table 1 presents a geospatial bivariate spreml analysis of the relationship of the CVIR to last month cannabis use, daily cannabis use, their interaction, cannabis quintiles and cannabis legal status. In each case these parameters are associated with (exponentiated) effect sizes of 1.2851, 1.2611, 0.9734, 2.2318 (Quintile 1 v 5) and 1.6063 (legal v illegal) respectively.

**Table 1.**
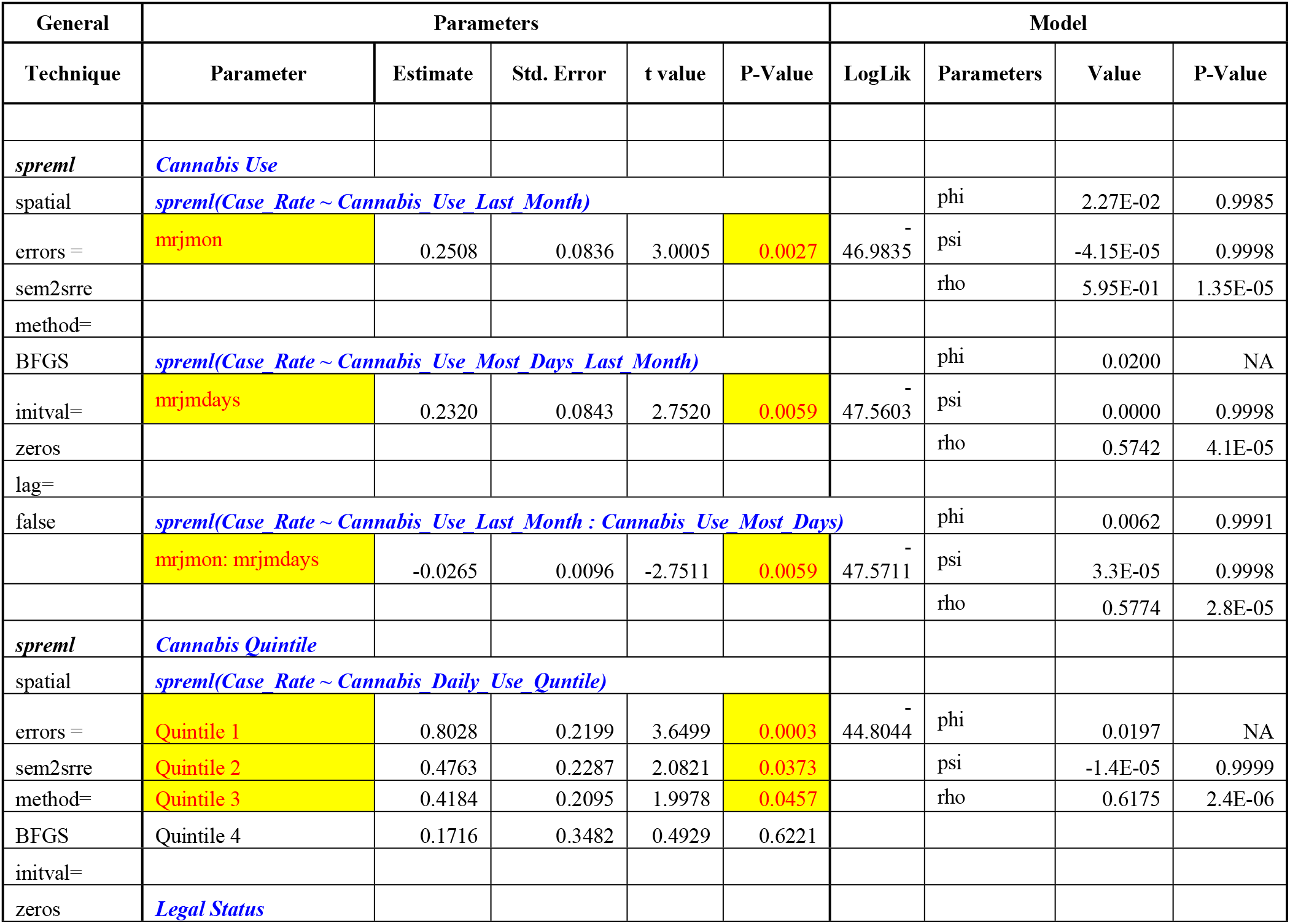
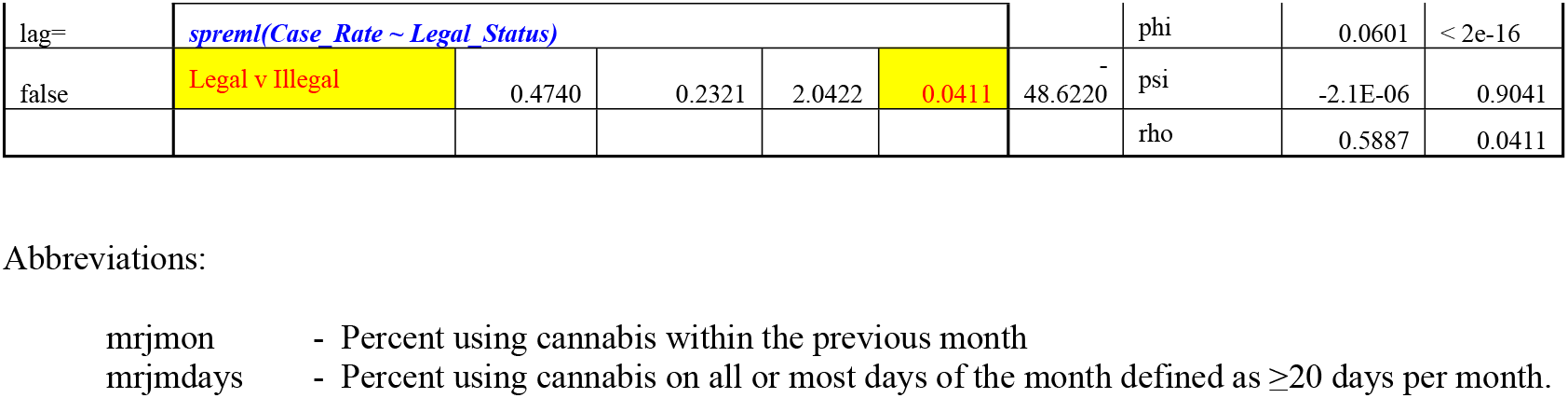
Bivariate Geospatial Regression Models of Cannabis Use

It was of interest to consider also the effect of the other variables each in their domain. Supplementary Table 3 presents the geospatial analysis of the data in the five domains of Flights, Median Household Income, Ethnicity, Population, and Drug Use. All three parameters in the flights domain are significant as might be expected. In this domain the Number of Flight Origins has the highest log likelihood ratio (logLik) so this is the parameter entered into full spatial models. Median household income is not significant. Two ethnicities are significant as noted. In the Population domain both total population and population density are significant.

The lower section of Supplementary Table 3 presents the Drug use domain. Three spreml models are presented each incorporating slightly different interaction structures between their terms as shown. Given that the final model has the highest log likelihood value (−32.4261) that is the model structure which is progressed to the full comprehensive models used subsequently. In this model the most significant term predictive of the CVIR is cannabis use (P=8.7×10^−7^). Cannabis use is included in 8 of the 11 terms remaining in the final model. The interaction between cannabis use last month and daily cannabis use is included in three terms and is also highly significant in its own right.

Given that many terms in the five domains of Supplementary Table 3 were significant it was of considerable interest to investigate how they compared when they were all combined together in a single comprehensive model. The results of additive models in all terms are shown in Supplementary Table 4. The number of flight origins and the population density are the remaining significant covariates after spgm model reduction. When the number of flights is used as the index of travel this term does not appear in the final model, but cannabis use persists as the most significant term (P=0.0079).

Table 2 presents final interactive spatial models after reduction via the spml, spgm and spreml algorithms. Interestingly travel, cannabis and opioid pain relievers are found to be significant in all final models. Terms including cannabis are most significant in the spml model, from P=7.3×10^−15^. Cannabis alone is significant (P=0.0365) in the spgm model, a technique which is sensitive to short panel datasets of this type. Cannabis is included in seven of nine terms, eight of twelve terms and seven of nine terms in the three models respectively. Interactions between last month cannabis use and daily cannabis use are included in three terms in each model. Tobacco, binge alcohol, cocaine and amphetamines did not appear in any final spatial models. Study of spreml model error structure confirmed that the full error structure (sem2srre without lagging) was indeed appropriate.

**Table 2.**
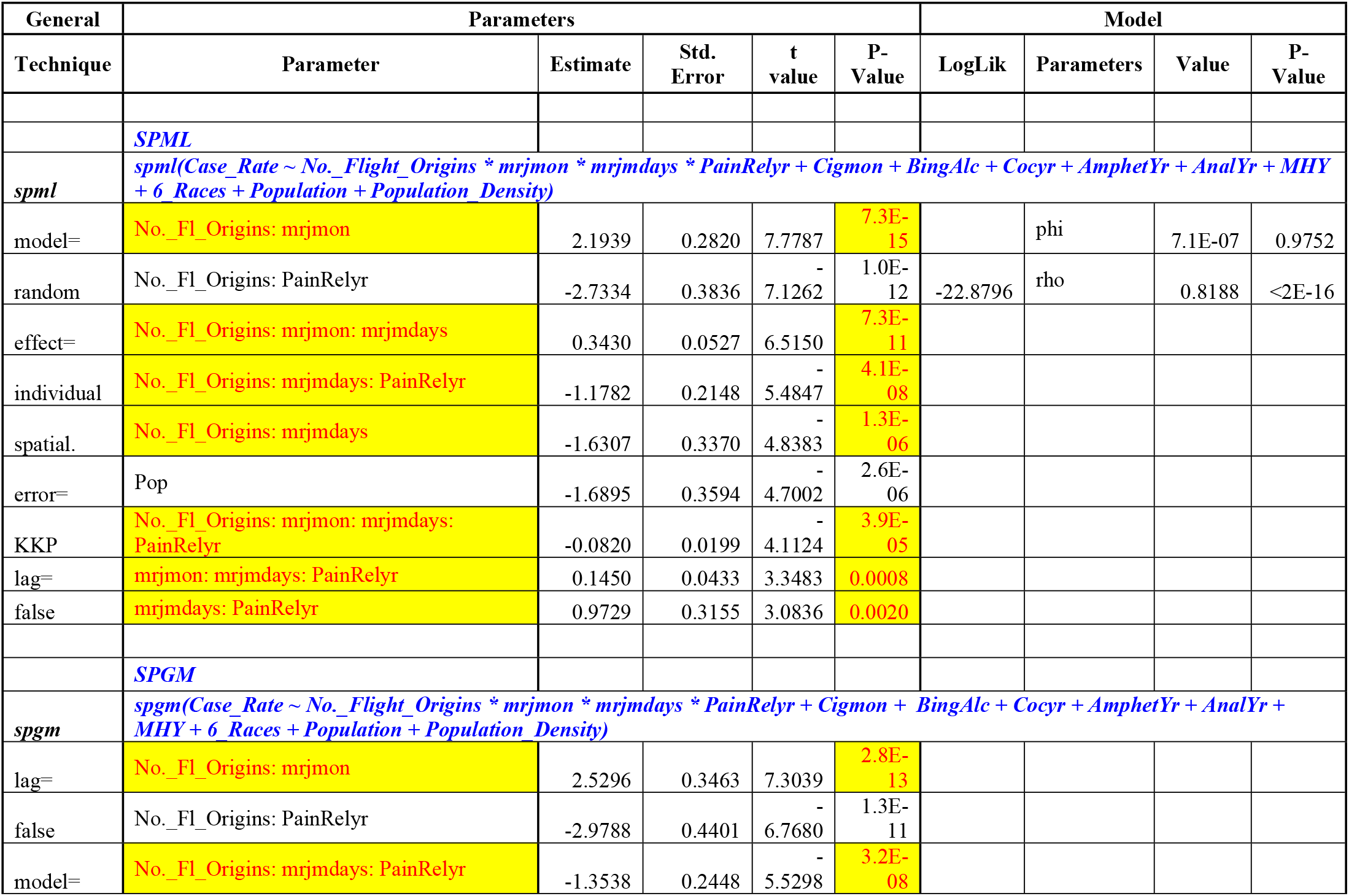
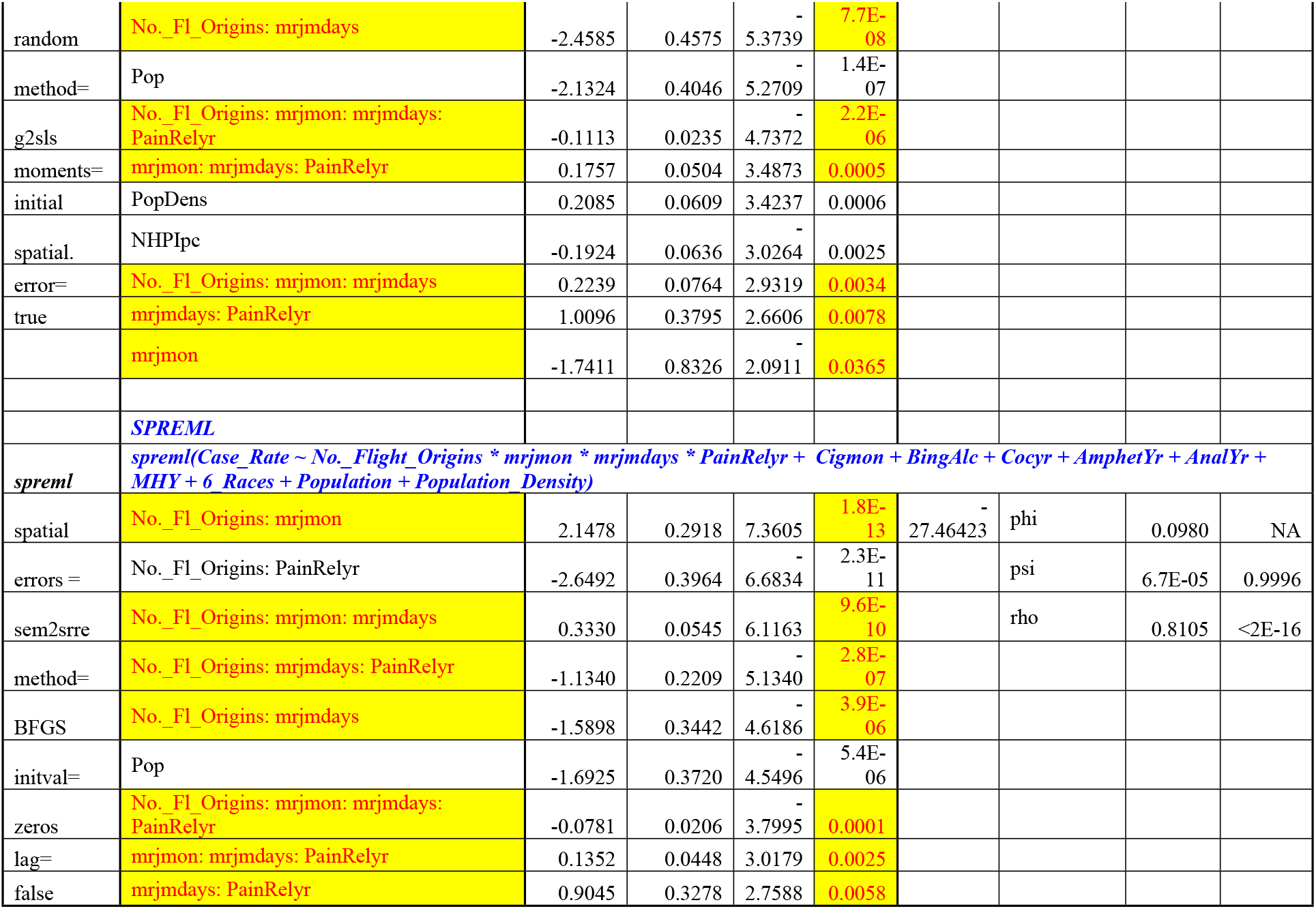
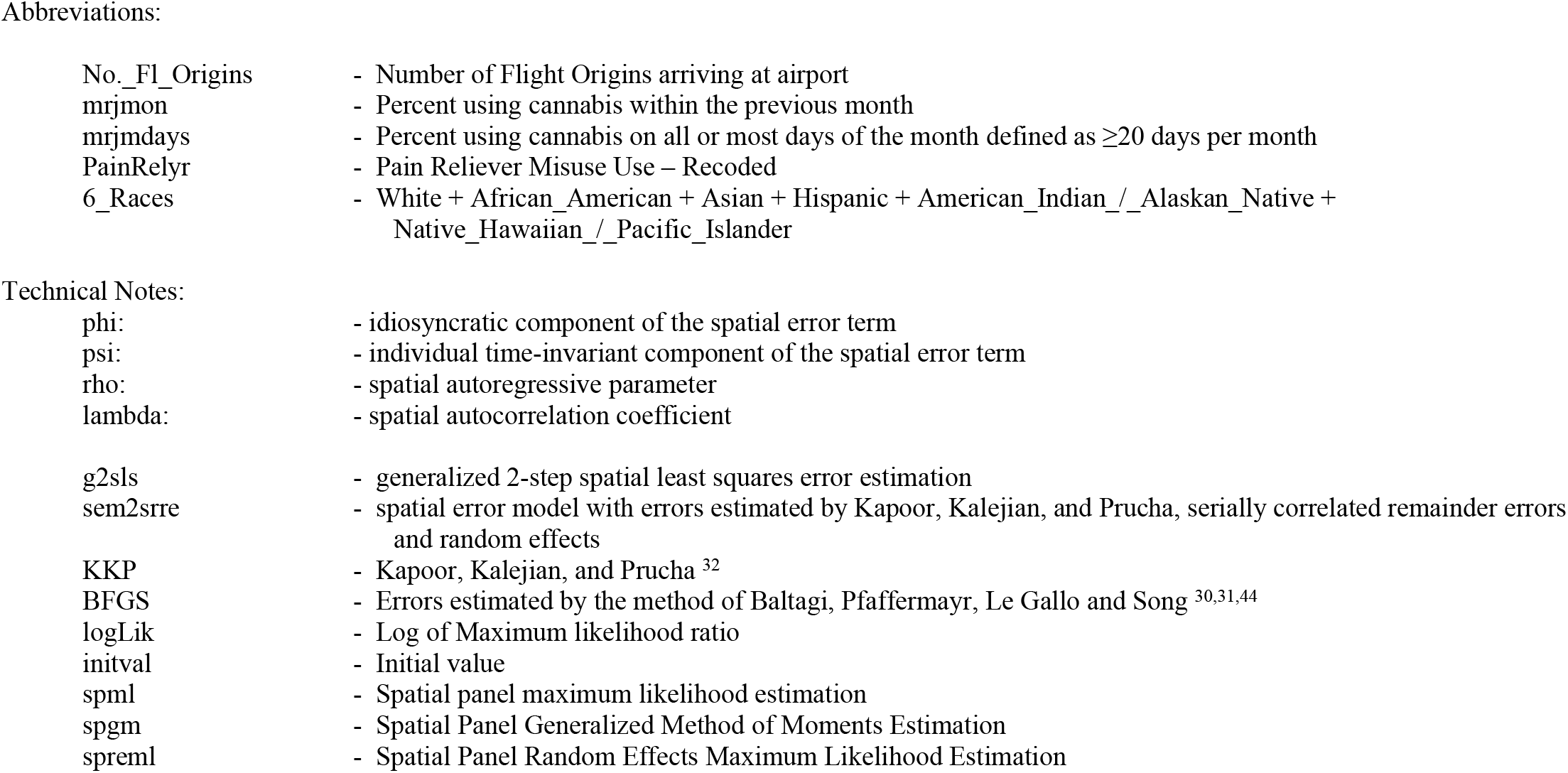
Final Geospatial Regression Models

## Discussion

Our study set out to explore the possible ecological and geospatial associations of cannabis use and coronavirus infection with the concern that cannabis-associated immunosuppression and cannabis contamination might exacerbate the global pandemic at a time when cannabis use and particularly the intensity of cannabis use is rising dramatically in many parts of USA and abroad. Bivariate evidence supported this hypothesis by demonstrating significant associations of daily cannabis use quintile with CVIR with the highest quintile having a prevalence ratio (PR, like odds ratio) of 5.11 (95%C.I. 4.90-5.33), an attributable fraction in the exposed of 80.45% (79.61-81.25%), and an attributable fraction in the population of 77.80% (76.88-78.68%) with a trend significant at P<10^−500^. Similarly when cannabis legalization was considered decriminalization was associated with an elevated CVIR prevalence ratio of 4.51 (95%C.I. 4.45-4.58), an attributable fraction in the exposed of 77.84% (77.50-78.17%) and an attributable fraction in the population of 51.22% (50.74-51.70%) and a trend significant at P<10^−500^. When the effect was studied in a multivariable geospatial model after controlling for international travel, ethnicity, income, population, population density and drug use interactive terms in last month cannabis were significant from 7.3×10^−15^ and daily cannabis use from 7.3×10^−11^. Cannabis use was independently predictive of CVIR in the final spgm model. These results strongly support the hypothesis of an ecological geospatial link between cannabis use and coronavirus infection rate.

Cannabinoids are known to interact with the immune system at multiple points including CB1 and CB2 receptors, six vanilloid channels, peroxisome proliferator-activated receptors (PPAR’s), serotonin, adenosine, histamine, glycine, sphingosine, dopamine and opioid receptors, three class A orphan G-protein coupled receptors (GPCR’s), toll-like receptors, T-cells, B-cells, macrophages and regulatory cells, effects on sodium channels and several types of potassium and calcium channels, modulation of GABA signalling and inhibition of cyclooxygenase and lipoxygenase enzymes, bind directly to mitochondria and cannabinoid receptors also form heterodimers with opioid, adenosine, dopamine, GABA and other GPCR’s and have myriad and major epigenetic effects ^13-20,33-38^.

The highly potent mammalian toxin carbofuran has also been described as being used on cannabis plants to prevent them being eaten by herbivores such as deer and has been found in cannabis plantations in large quantities ^24^. This extremely potent toxin is an acetylcholinesterase inhibitor banned in USA in 1991 in granular form and in liquid form in 2009 by the Environmental Protection Agency (EPA) after it was implicated in the death of over 1,000,000 birds including eagles in USA and many lions in Africa ^39,40^. Concerns relating to carbofuran contamination of groundwater and fresh drinking water supplies have been expressed by the US EPA and WHO ^41-43^.

To our knowledge this investigation is the first report of a positive association between CVIR and cannabis. Nevertheless given that cannabis is known to have significant immunosuppressive effects by many biological mechanisms, and that reports of contamination of cannabis with diverse chemical, microbial and fungal organisms are not uncommon ^22-25^, and given the very high levels of statistical significance demonstrated in the present analysis by several techniques, we are concerned that this effect is likely robust and generalizable. In the context of rapidly accelerating pandemics of both cannabis and coronavirus this suggests a biological and mechanistic synergism which is of considerable concern. An interesting issue raised by this data is that cannabis-related -immunosuppression and -contamination is likely reversible upon cessation of exposure. This is an important issue requiring further research.

This report has several strengths and limitations. Our study is timely, and uses a current dataset for CVIR. The study uses a well validated nationally representative drug use dataset, which is widely studied and extensively quoted. Importantly we use two metrics of cannabis use including one which provides a measure of daily (or near daily) cannabis use, which has been shown to be the major parameter of American cannabis consumption ^27^. We use a very large dataset of international flight arrivals into USA which captures the whole population of these events over a 12 month period. US Census Bureau data is used to source state population, income and ethnicity data from the well validated American Community Survey. Our analysis reaches similar conclusions by several different pathways in both bivariate and multivariable analyses. There is good concordance between models utilizing the spml, spgm and spreml geospatial algorithms. All our major results are at very high levels of statistical significance. The limitations of our study relate to its uncontrolled design. Case control studies cannot be considered in such situations since it is unethical to expose patients to a real risk of mortality in the absence of definitive treatment or vaccination (at the time of writing). Moreover our results are spatially restricted to state level data. For example upstate New York is very rural, but Manhattan is one of the most densely populated places on the planet.

The broader geospatial level of our study was not able to capture such important details which are likely of particular importance to socially transmissible agents such as coronavirus. Further studies at higher geospatial levels of resolution are strongly and urgently indicated.

In summary we found strong bivariate and multivariable confirmatory evidence for the hypothesis that cannabis use is associated with coronavirus infection. A strong quintile effect was noted along with a prominent effect of cannabis decriminalization. After adjustment cannabis use emerged as a persistent, independent and robust correlate of CVIR at high levels of significance. This finding is of concern and suggests a powerful negative feedforward interaction between two major public health challenges faced by USA and the international community. Given the immediate salience and potent imminence of the coronavirus epidemic this association is well worth further immediate epidemiological research. The present report indicates stricter cannabis controls as one public health measure by which to address an infectious challenge and support cellular and soluble immunity for the whole community.

## Data Availability

Study data is made available with this paper in the online supplementary material.

## Acknowledgements

All authors had full access to all the data in the study and takes responsibility for the integrity of the data and the accuracy of the data analysis.

## Authorship Contributions

ASR assembled the data, designed and conducted the analyses, and wrote the first manuscript draft. GKH provided technical and logistic support, co-wrote the paper, assisted with gaining ethical approval, provided advice on manuscript preparation and general guidance to study conduct.

---

**Supplementary Table 1.** Cannabis Quintiles & Legal Status Designations

| State | Quintile Daily Cannabis Use | Legal Status 2020 |
| --- | --- | --- |
| Alabama | Quintile 3 | Illegal |
| Alaska | Quintile 5 | Legal |
| Arizona | Quintile 2 | Illegal |
| Arkansas | Quintile 4 | Illegal |
| California | Quintile 1 | Legal |
| Colorado | Quintile 1 | Legal |
| Connecticut | Quintile 3 | Decriminalized |
| Delaware | Quintile 5 | Decriminalized |
| Florida | Quintile 1 | Illegal |
| Georgia | Quintile 2 | Illegal |
| Hawaii | Quintile 5 | Decriminalized |
| Idaho | Quintile 5 | Illegal |
| Illinois | Quintile 1 | Legal |
| Indiana | Quintile 2 | Illegal |
| Iowa | Quintile 5 | Illegal |
| Kansas | Quintile 5 | Illegal |
| Kentucky | Quintile 3 | Illegal |
| Louisiana | Quintile 3 | Illegal |
| Maine | Quintile 3 | Legal |
| Maryland | Quintile 3 | Decriminalized |
| Massachusetts | Quintile 2 | Legal |
| Michigan | Quintile 1 | Legal |
| Minnesota | Quintile 3 | Decriminalized |
| Mississippi | Quintile 4 | Decriminalized |
| Missouri | Quintile 3 | Illegal |
| Montana | Quintile 5 | Illegal |
| Nebraska | Quintile 5 | Decriminalized |
| Nevada | Quintile 3 | Legal |
| New Hampshire | Quintile 5 | Decriminalized |
| New Jersey | Quintile 2 | Illegal |
| New Mexico | Quintile 4 | Decriminalized |
| New York | Quintile 1 | Decriminalized |
| North Carolina | Quintile 2 | Decriminalized |
| North Dakota | Quintile 5 | Decriminalized |
| Ohio | Quintile 1 | Decriminalized |
| Oklahoma | Quintile 4 | Illegal |
| Oregon | Quintile 2 | Legal |
| Pennsylvania | Quintile 1 | Illegal |
| Rhode Island | Quintile 5 | Decriminalized |
| South Carolina | Quintile 3 | Illegal |
| South Dakota | Quintile 5 | Illegal |
| Tennessee | Quintile 2 | Illegal |
| Texas | Quintile 1 | Illegal |
| Utah | Quintile 5 | Illegal |
| Vermont | Quintile 5 | Legal |
| Virginia | Quintile 2 | Illegal |
| Washington | Quintile 1 | Legal |
| West Virginia | Quintile 5 | Illegal |
| Wisconsin | Quintile 2 | Illegal |
| Wyoming | Quintile 5 | Illegal |

**Supplementary Table 2.** Quintile & Legal Status Analysis

| Group | Cases | Controls | Case_Rate<br>/ 10,000 | Prevalence Ratio | Attributable<br>Fraction in<br>Exposed (%) | Attributable<br>Fraction in<br>Population (%) |
| --- | --- | --- | --- | --- | --- | --- |
| **Quintile** |  |  |  |  |  |  |
| Quintile 5 | 2,263 | 23,881,293 | 9.5 | - | - | - |
| Quintile 4 | 1,388 | 11,988,616 | 11.6 | 1.22 (1.14-1.31) | 18.15 (12.49-23.44) | 6.90 (4.51-9.24) |
| Quintile 3 | 7,342 | 44,375,104 | 16.5 | 1.75 (1.67-1.83) | 42.72 (39.96-45.36) | 32.66 (30.19-35.04) |
| Quintile 2 | 16,219 | 74,658,238 | 21.7 | 2.29 (2.19-2.40) | 56.38 (54.41-58.25) | 47.48 (47.48-51.39) |
| Quintile 1 | 66,531 | 167,221,538 | 39.8 | 5.11 (4.90-5.33) | 80.45 (79.61-81.25) | 77.80 (76.88-78.68) |
| **Legal Status** |  |  |  |  |  |  |
| Illegal | 25,943 | 161,869,909 | 1.60 | - | - | - |
| Legal | 17,878 | 91,266,635 | 1.96 | 1.22 (1.20-1.25) | 18.18 (16.61-19.72) | 7.42 (6.69-8.13) |
| Decriminalized | 49,922 | 68,988,245 | 7.24 | 4.51 (4.45-4.58) | 77.84 (77.50-78.17) | 51.22 (50.74-51.70) |

**Supplementary Table 3.**
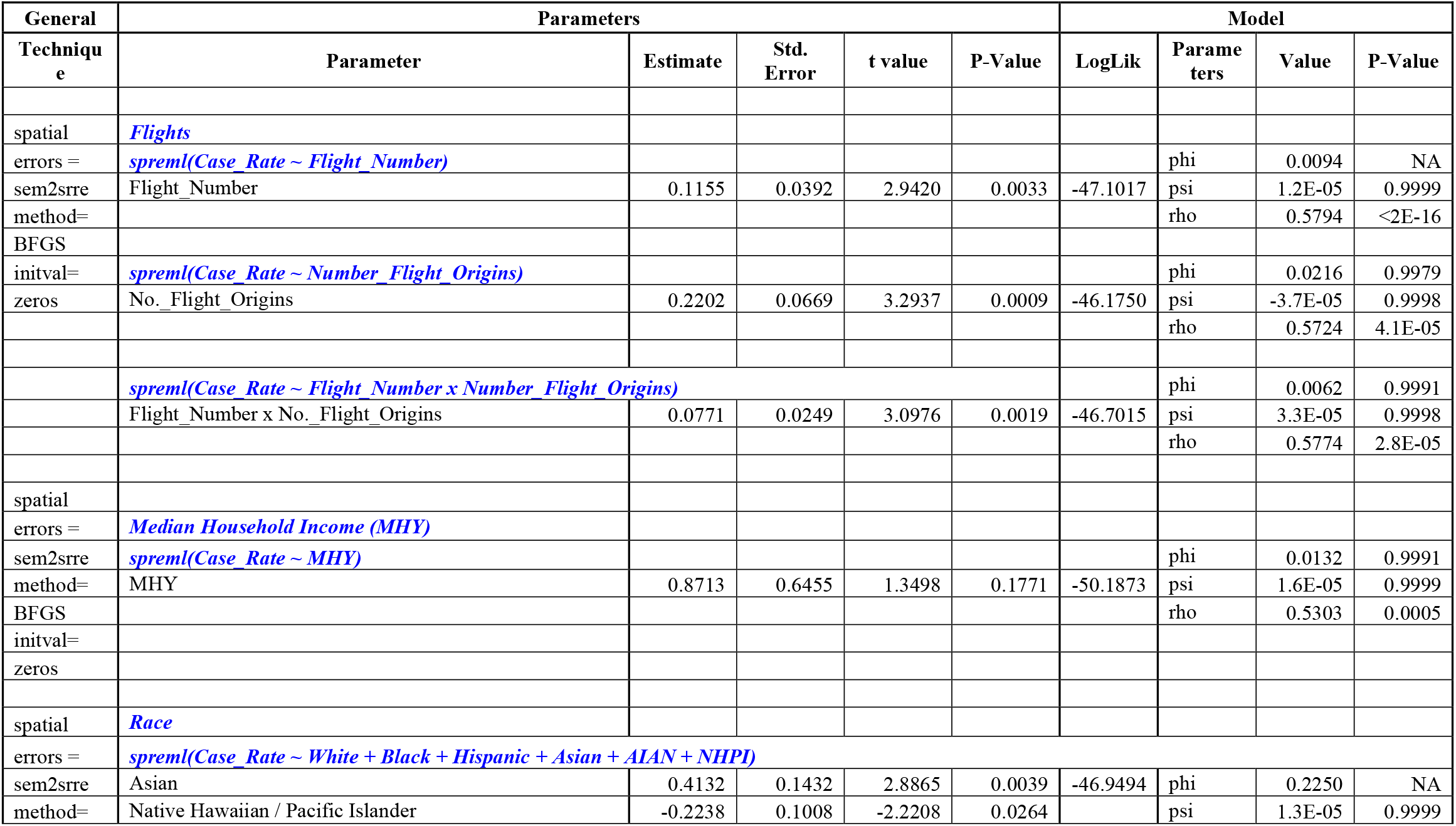
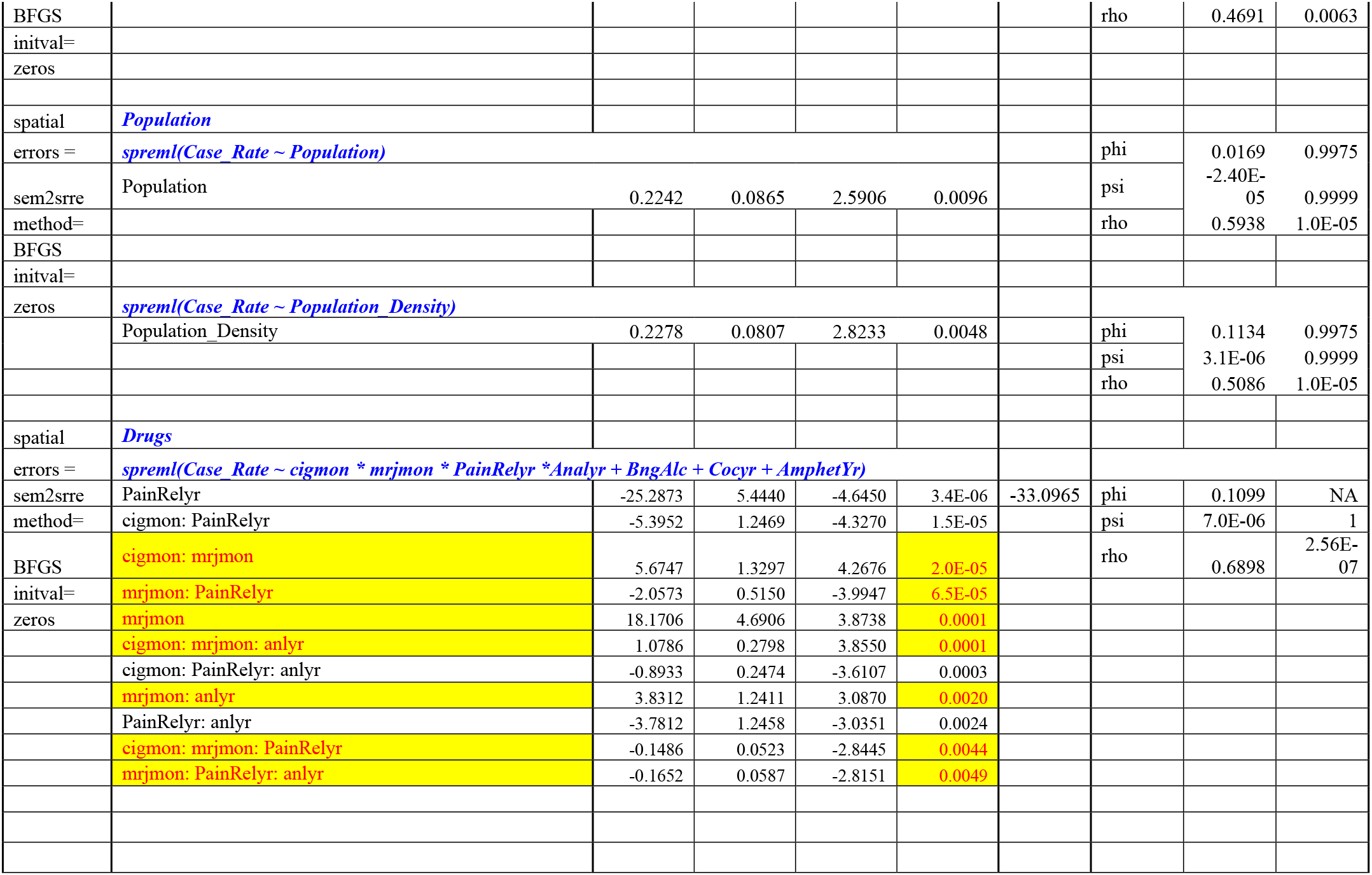
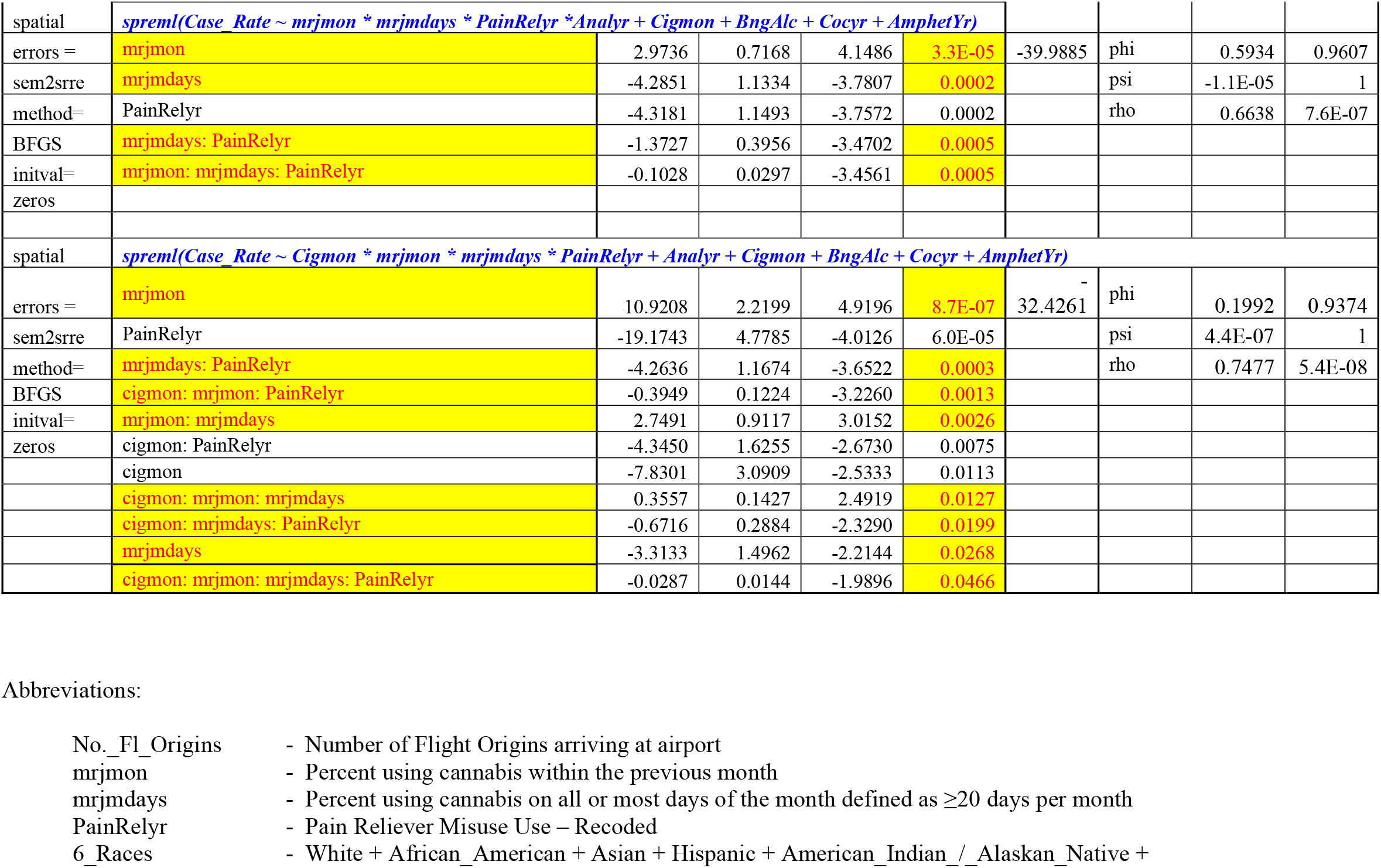
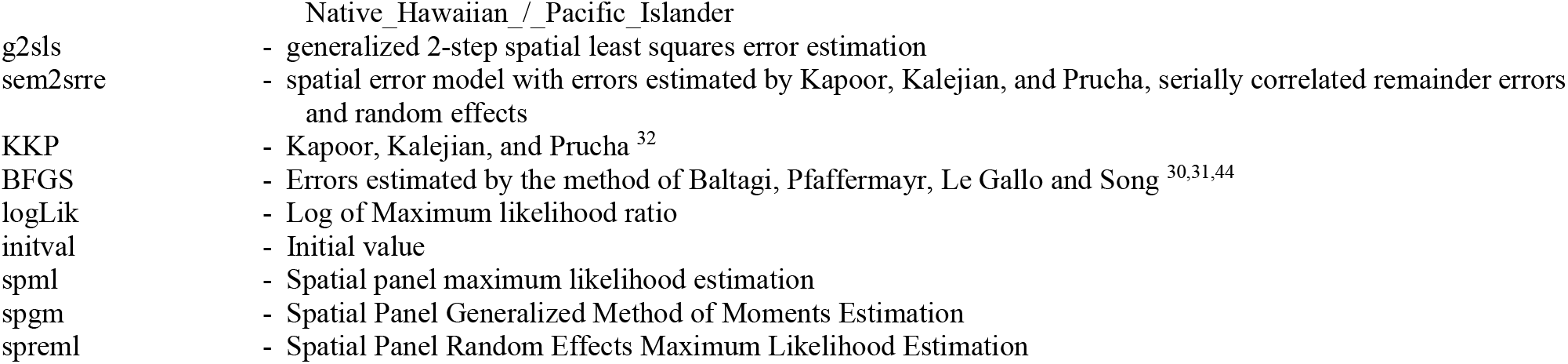
Geospatial Spreml Regression on Single Group Variables

**Supplementary Table 4.**
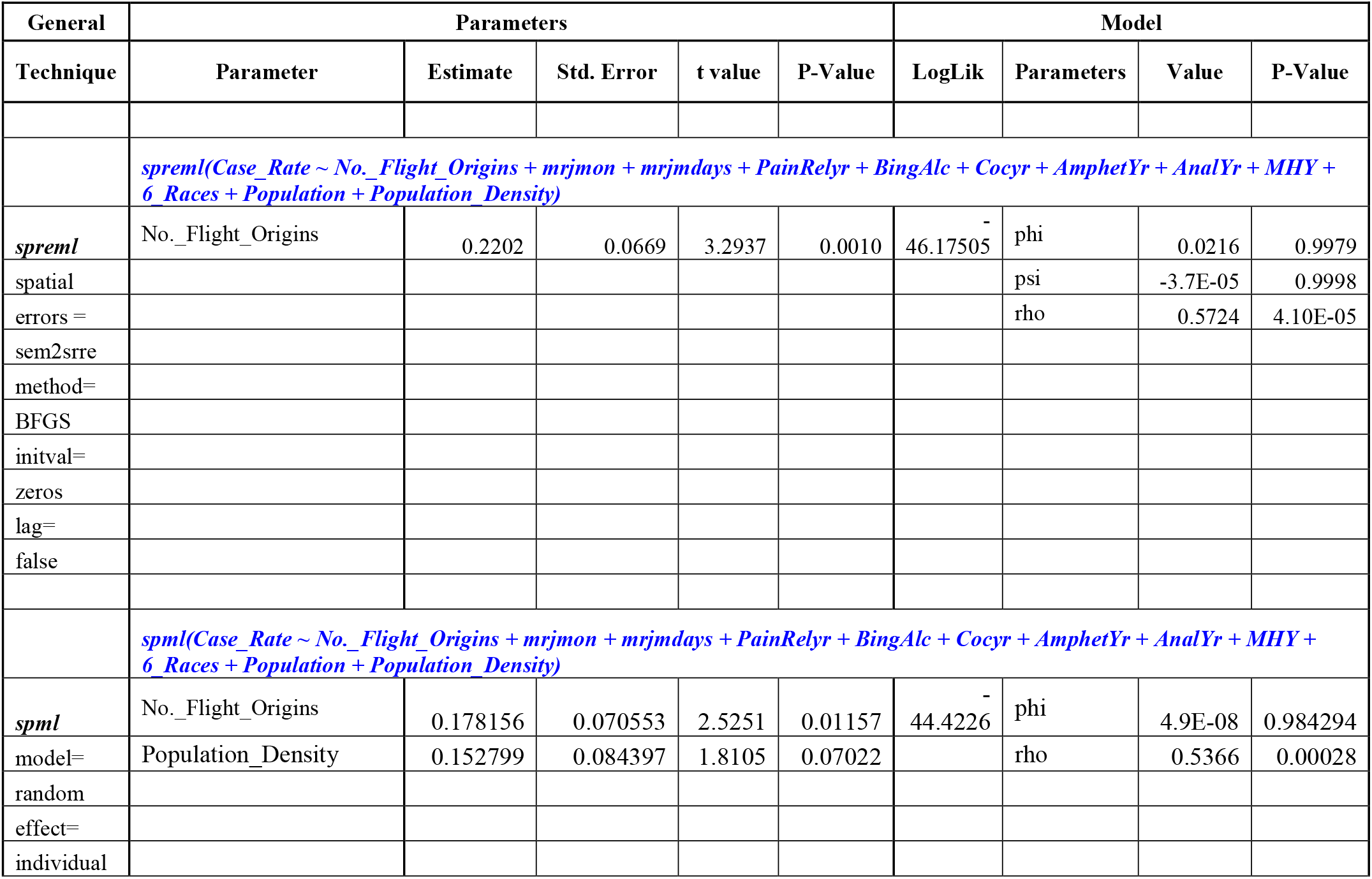
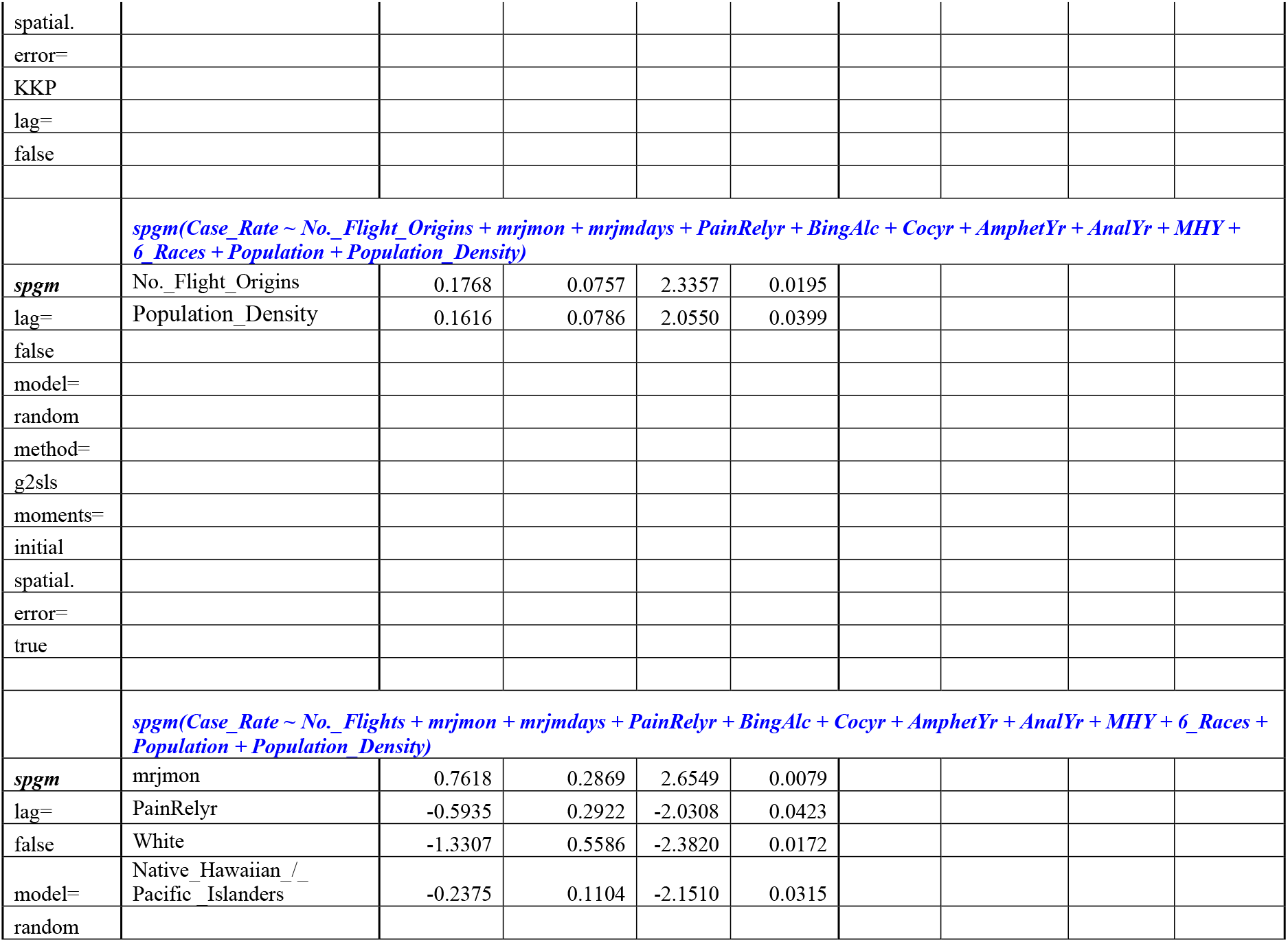
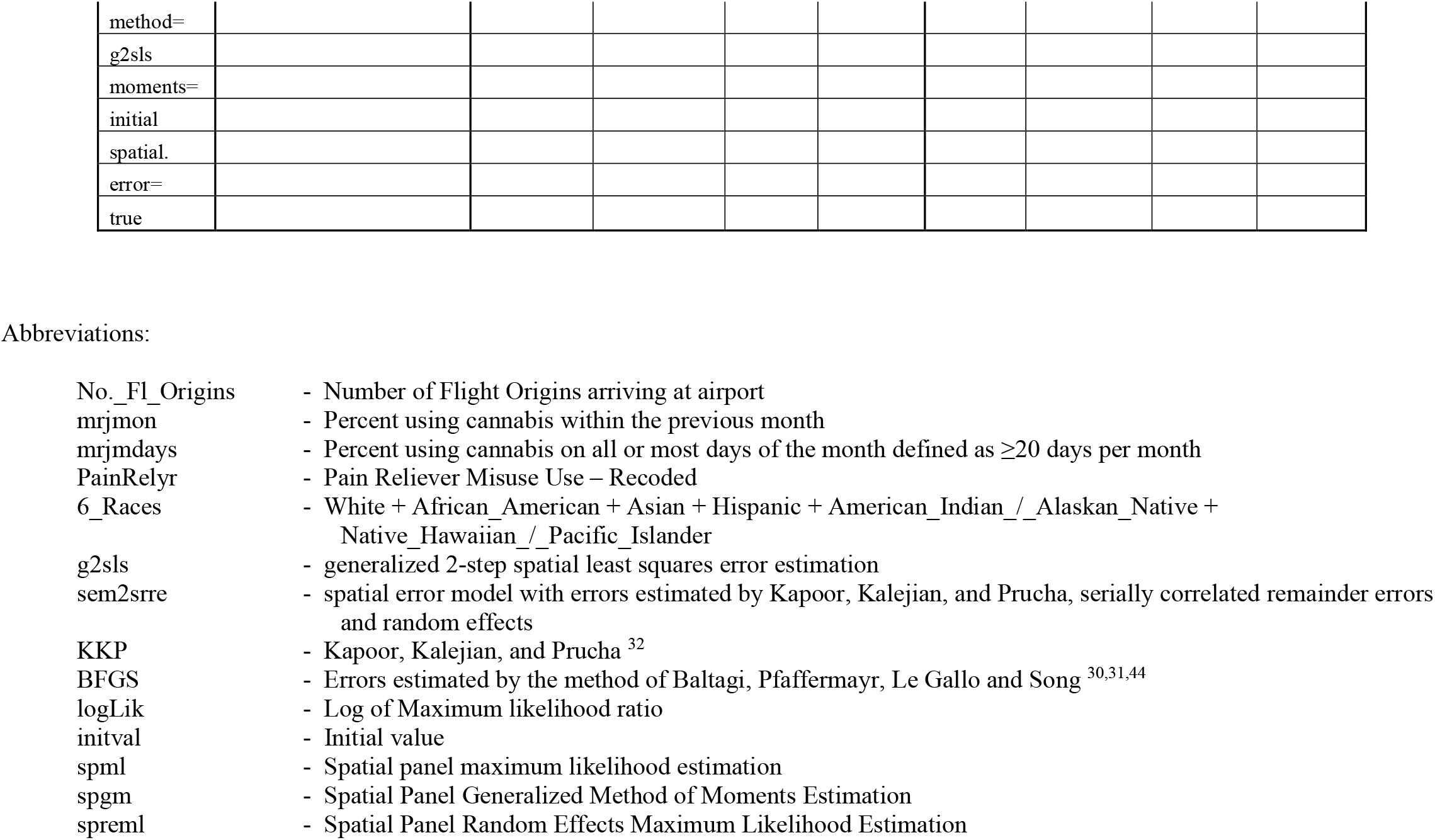
Additive Geospatial Regressions

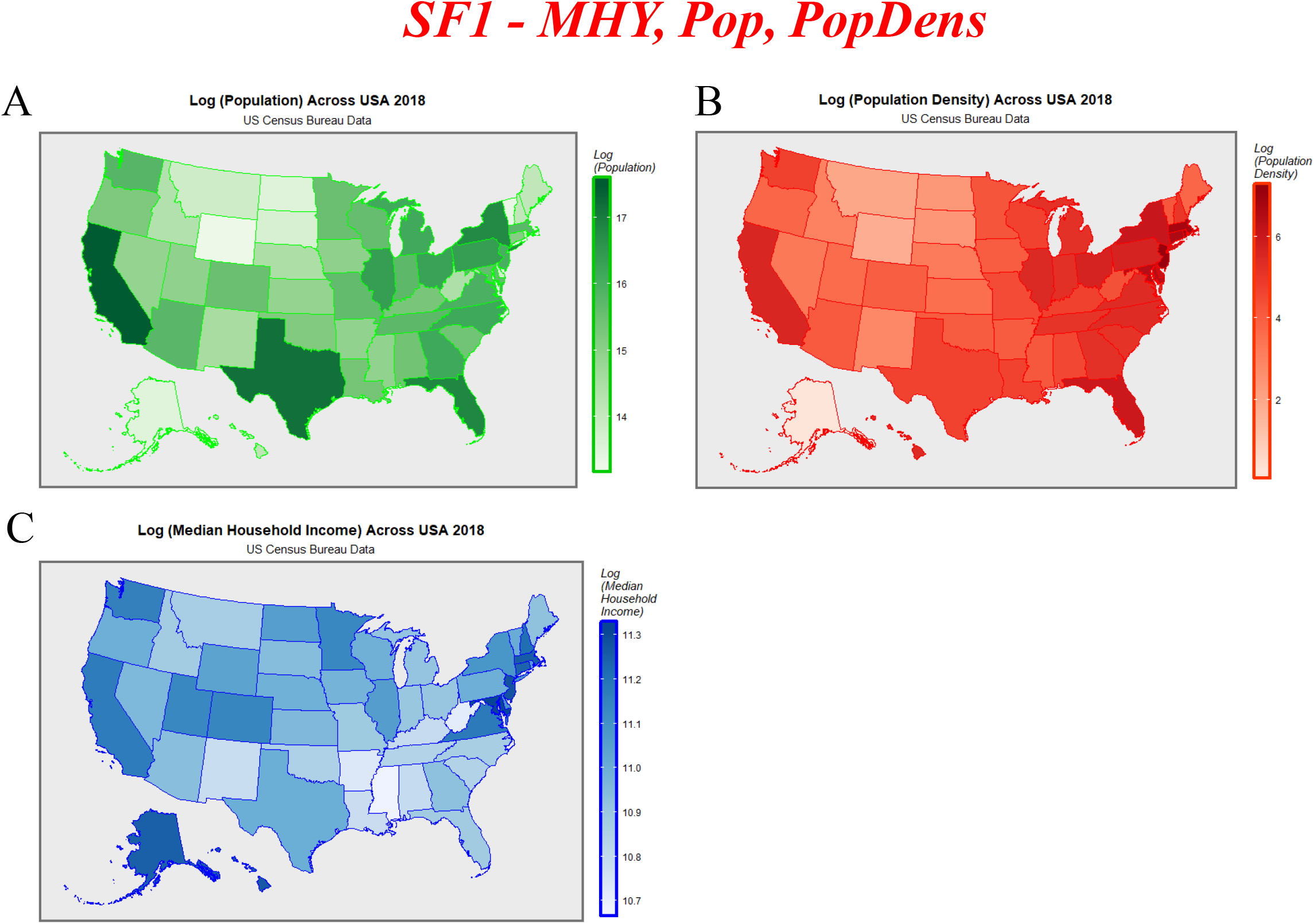

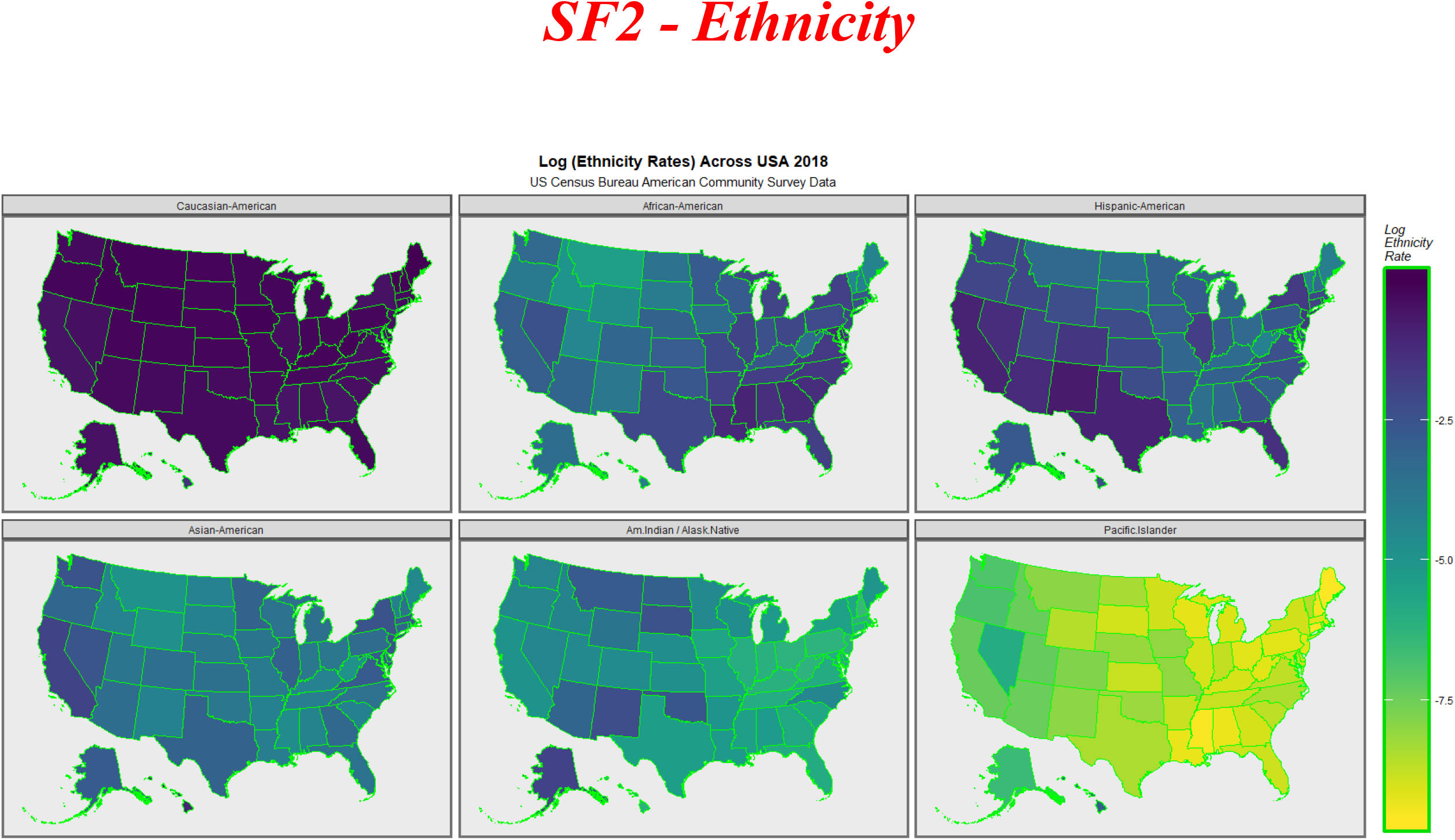

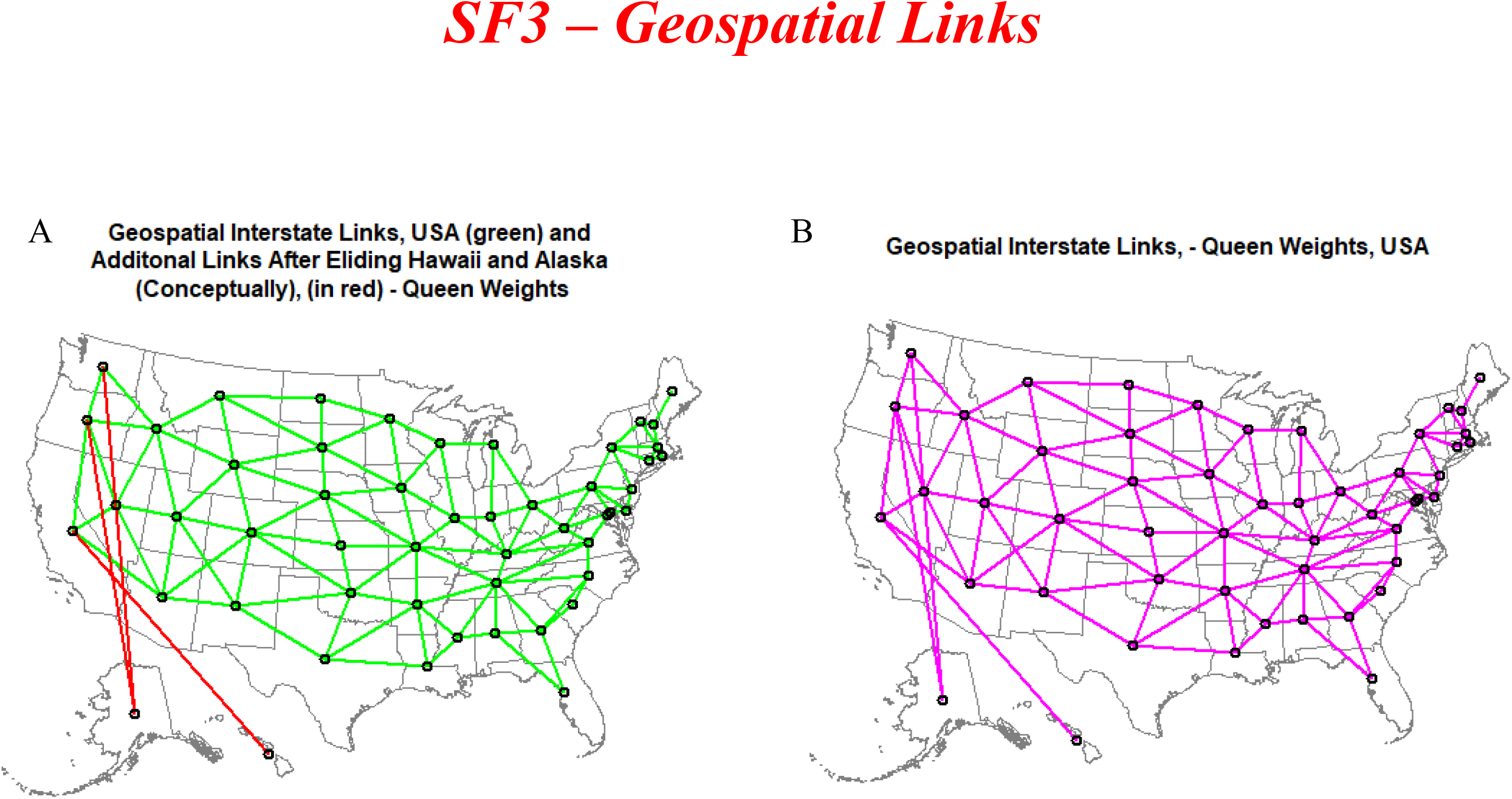

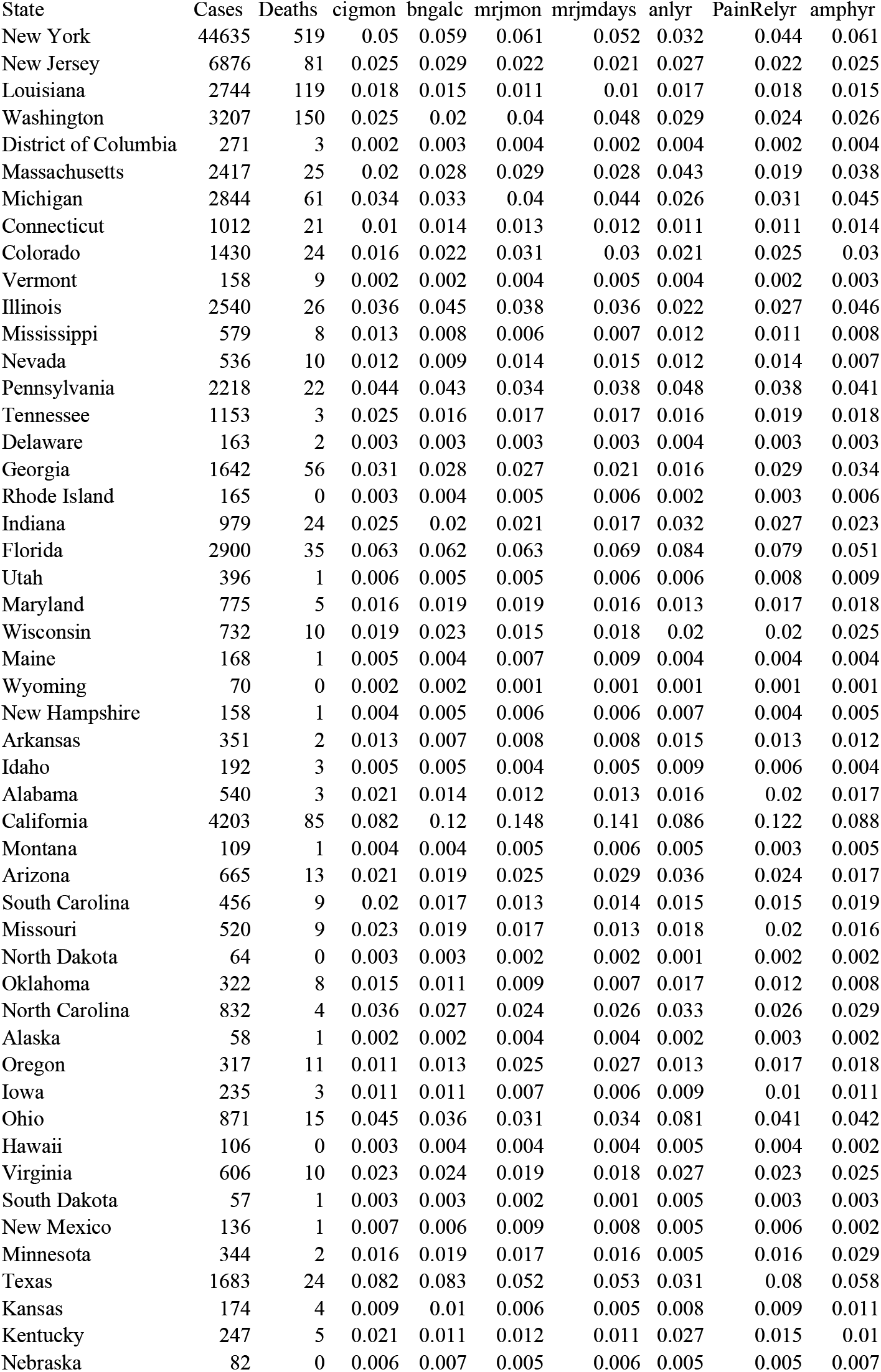
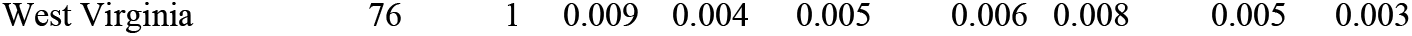
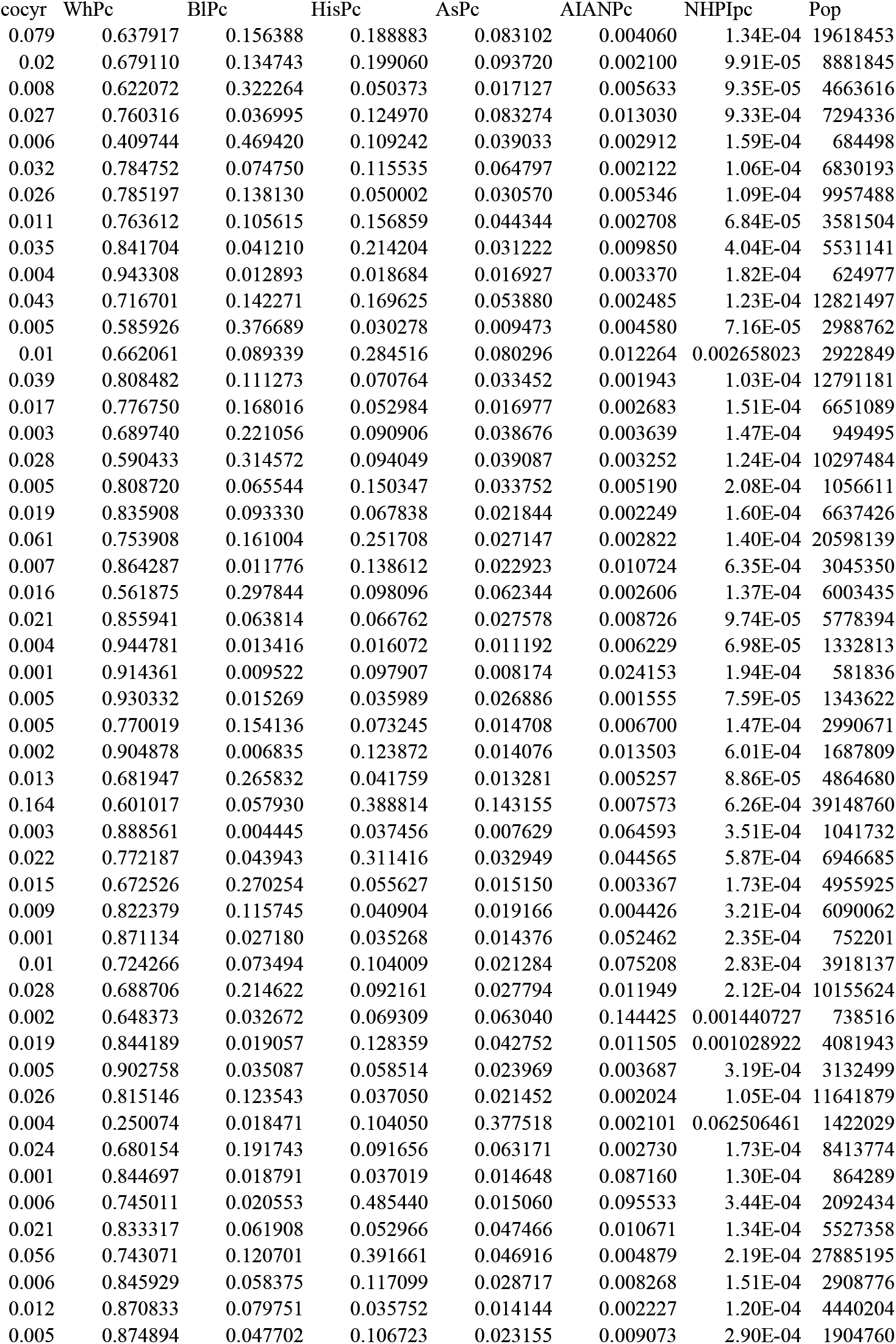
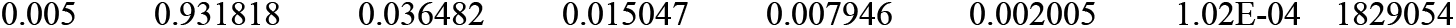
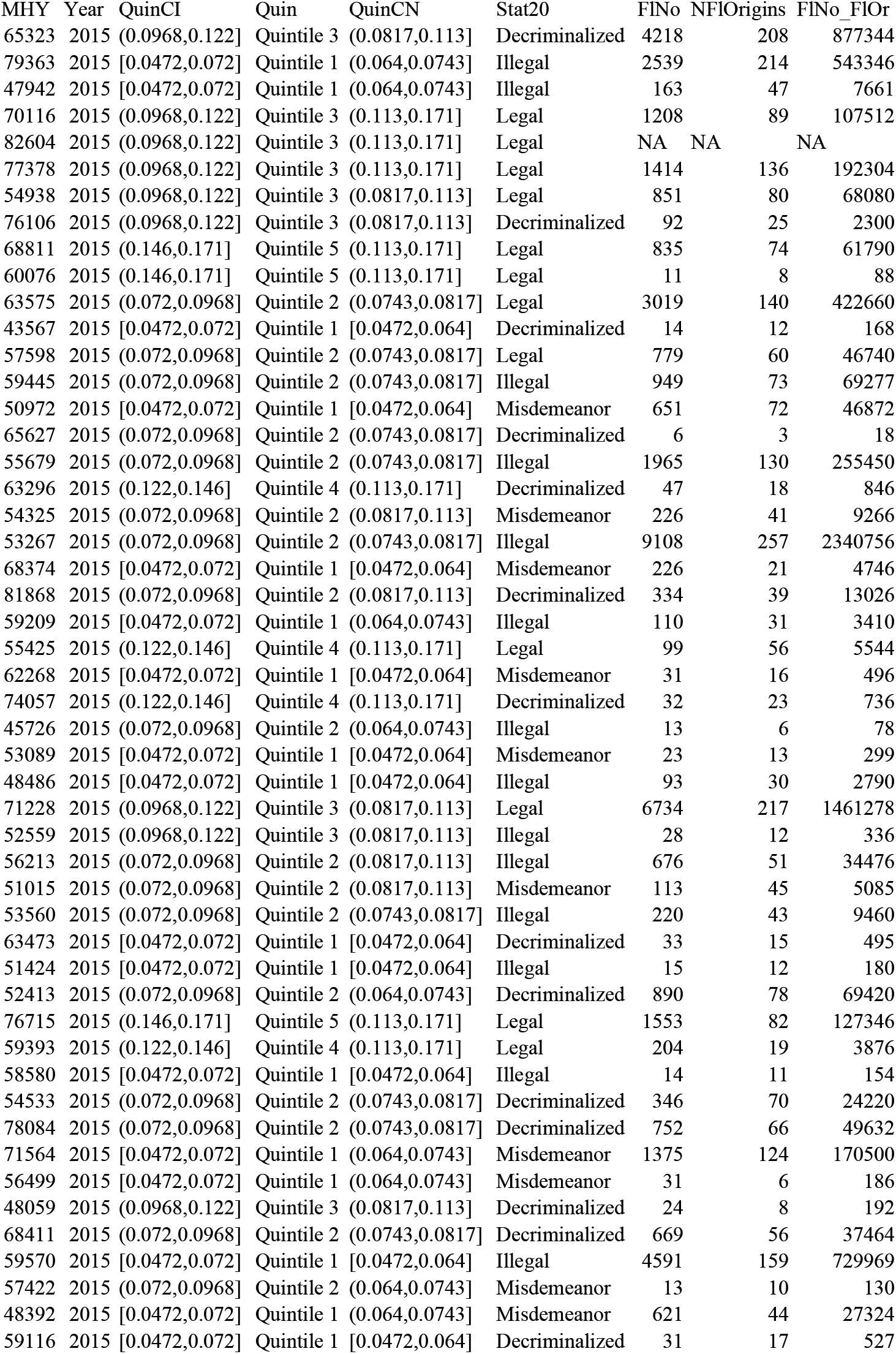
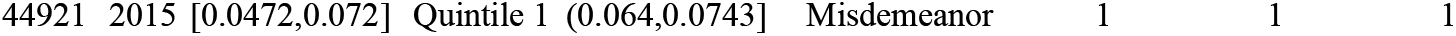
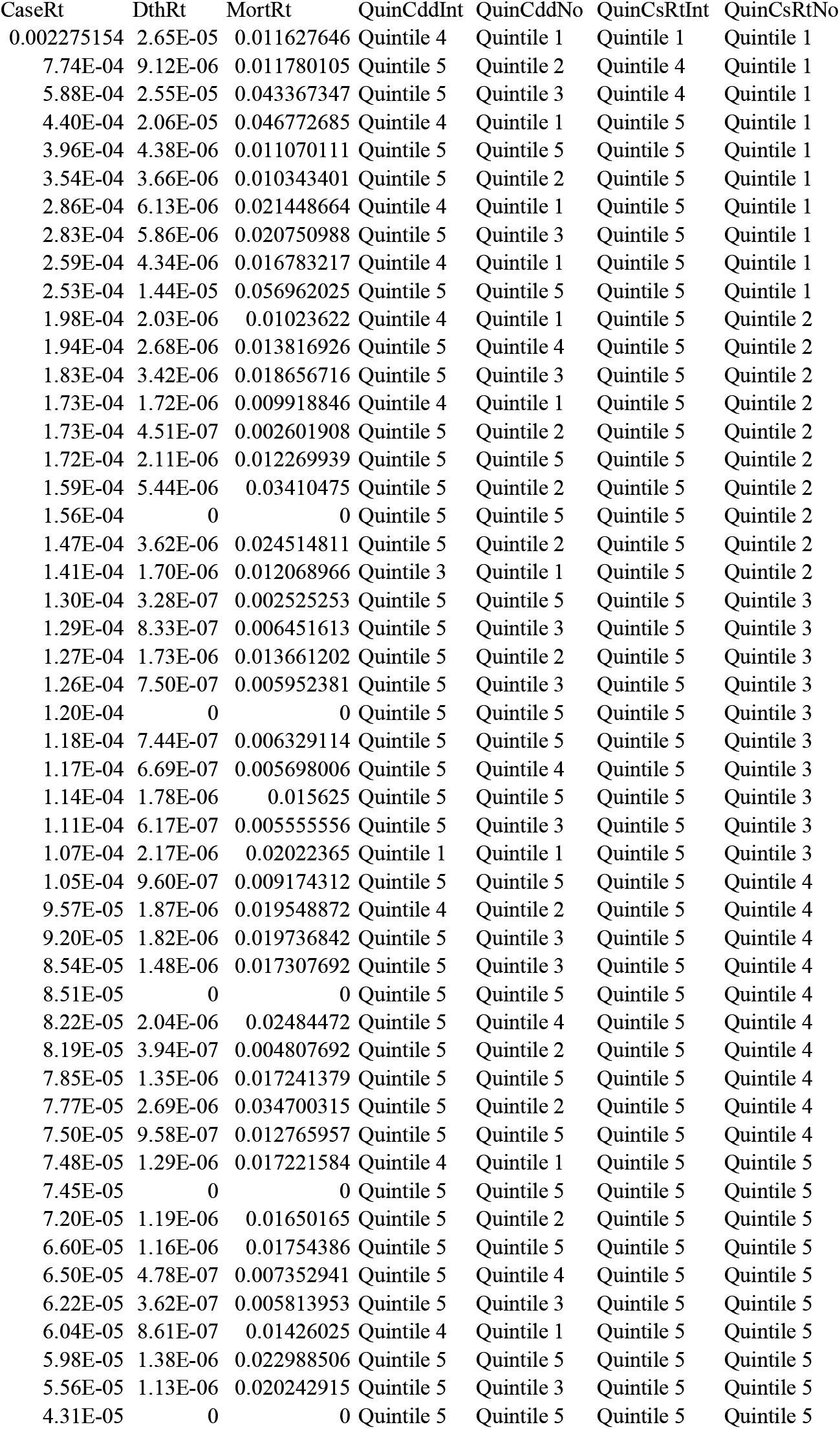
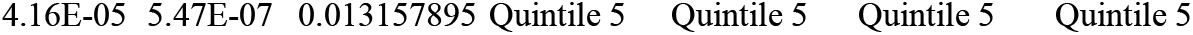
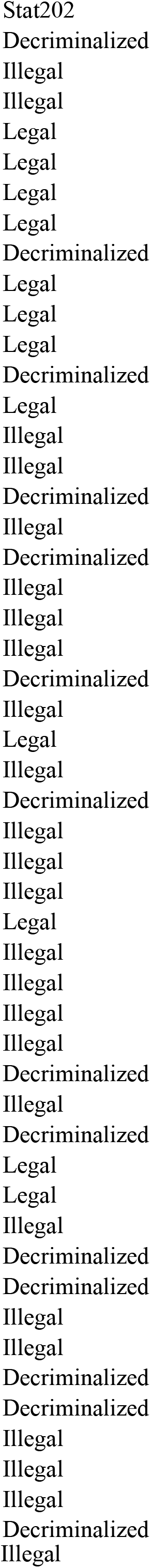
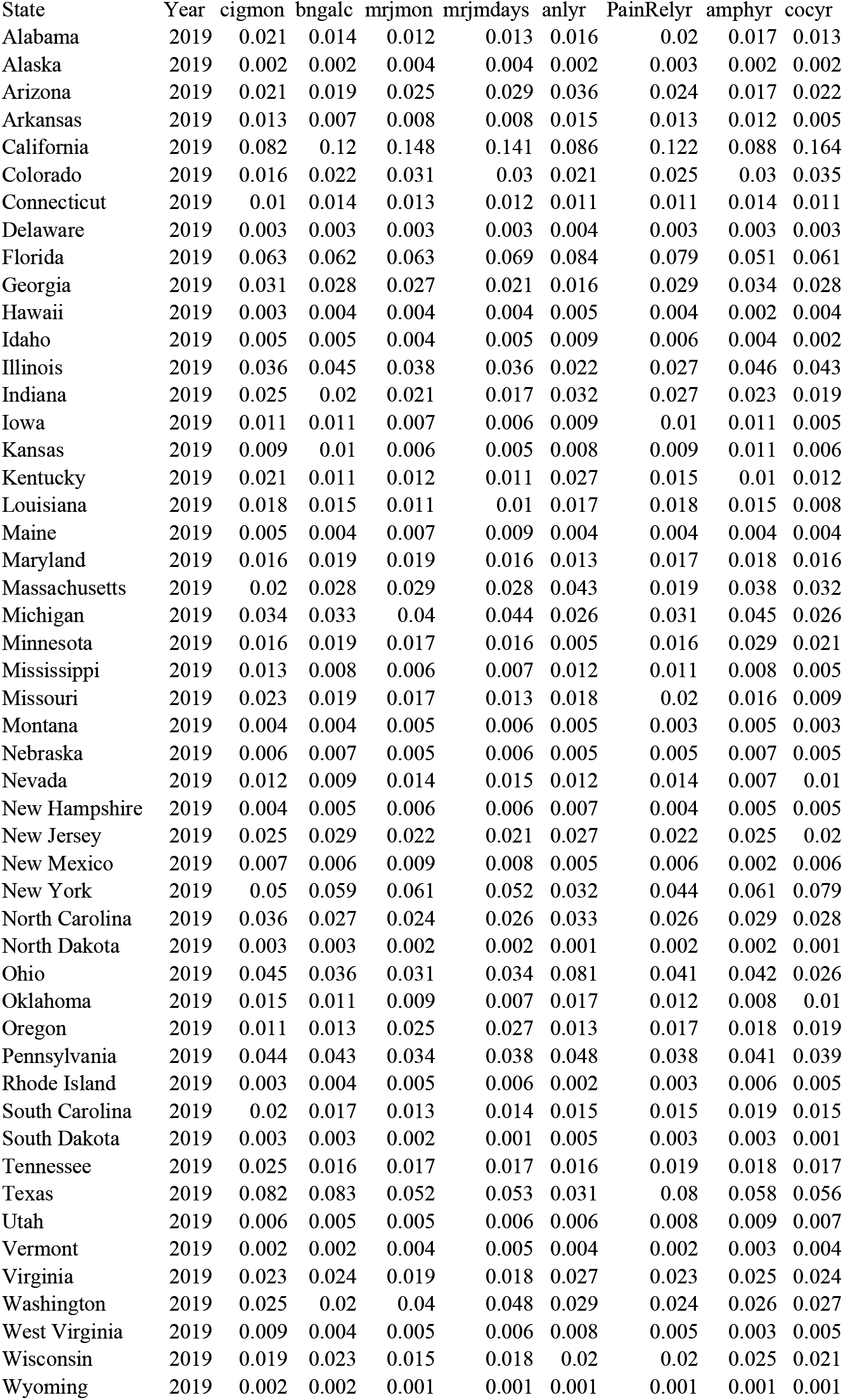
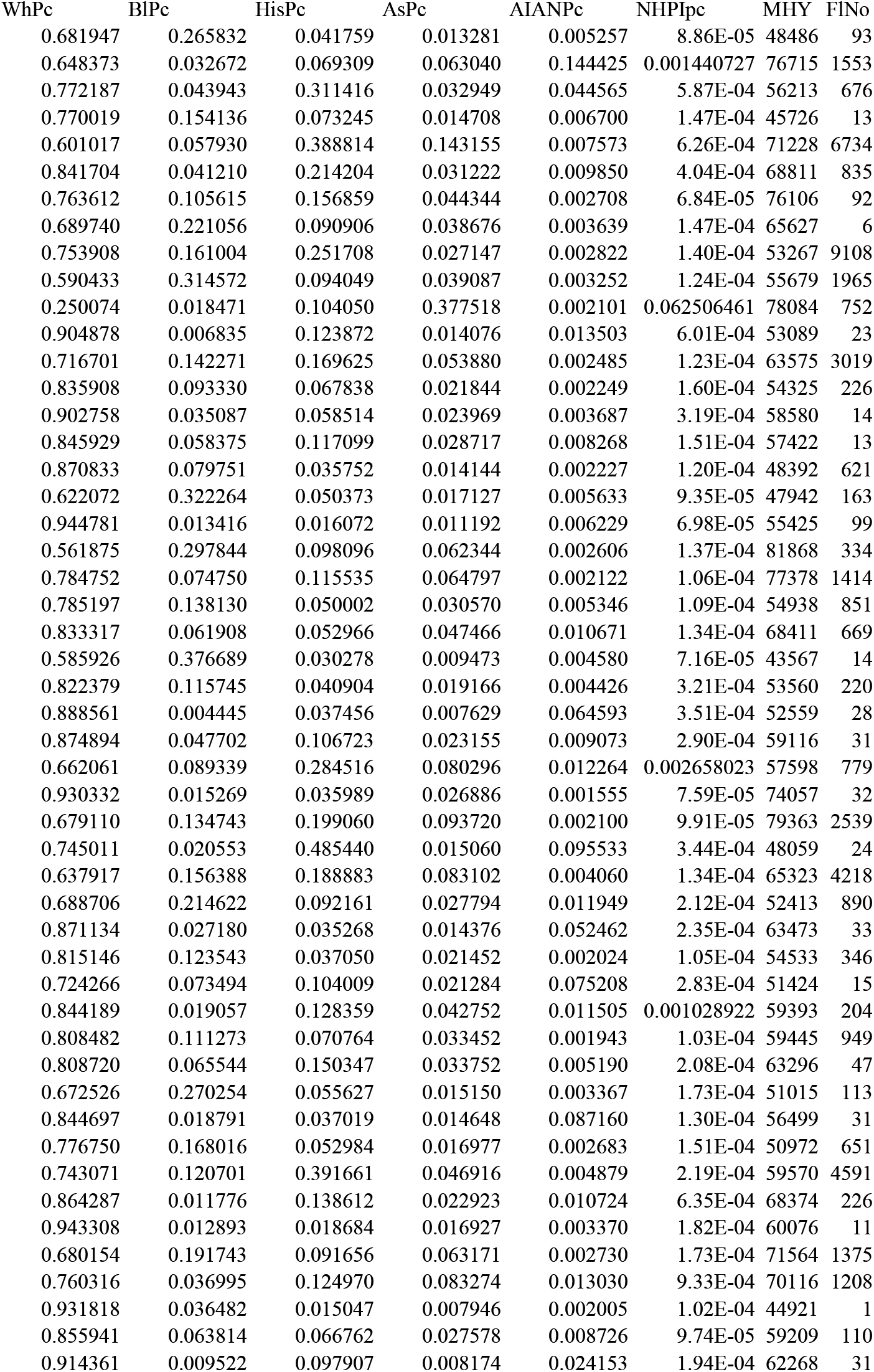
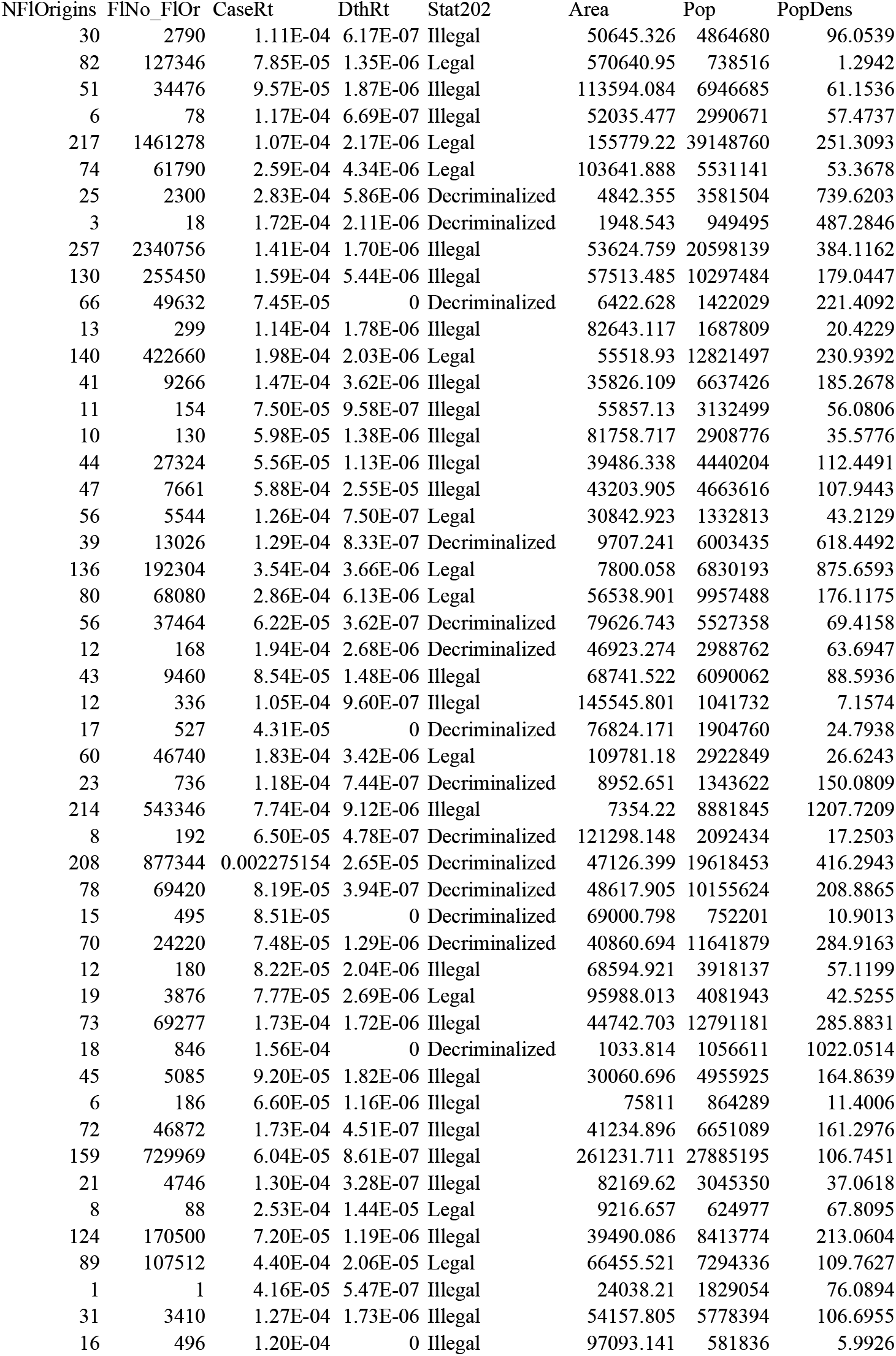

## STROBE Statement—checklist of items that should be included in reports of observational studies

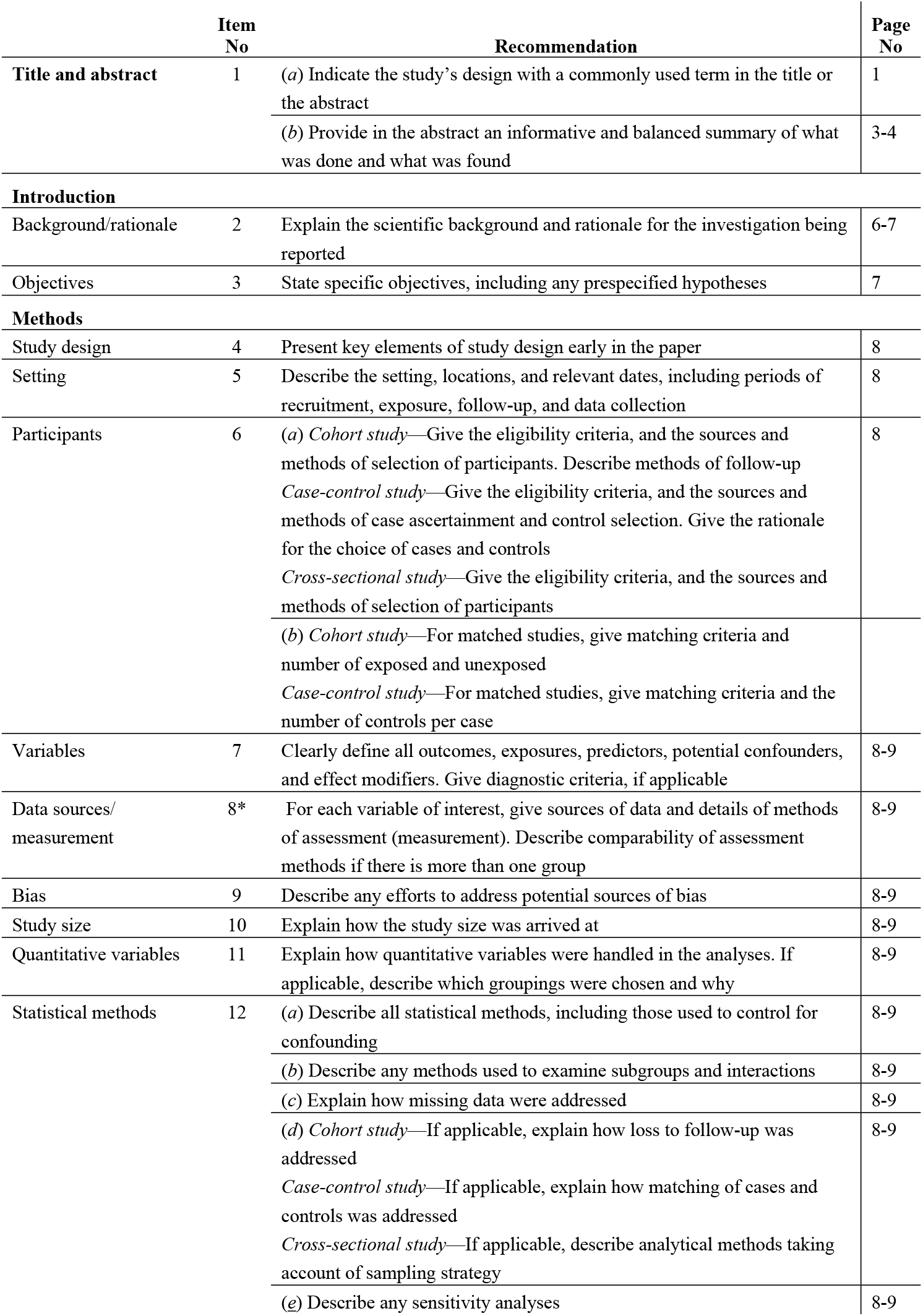
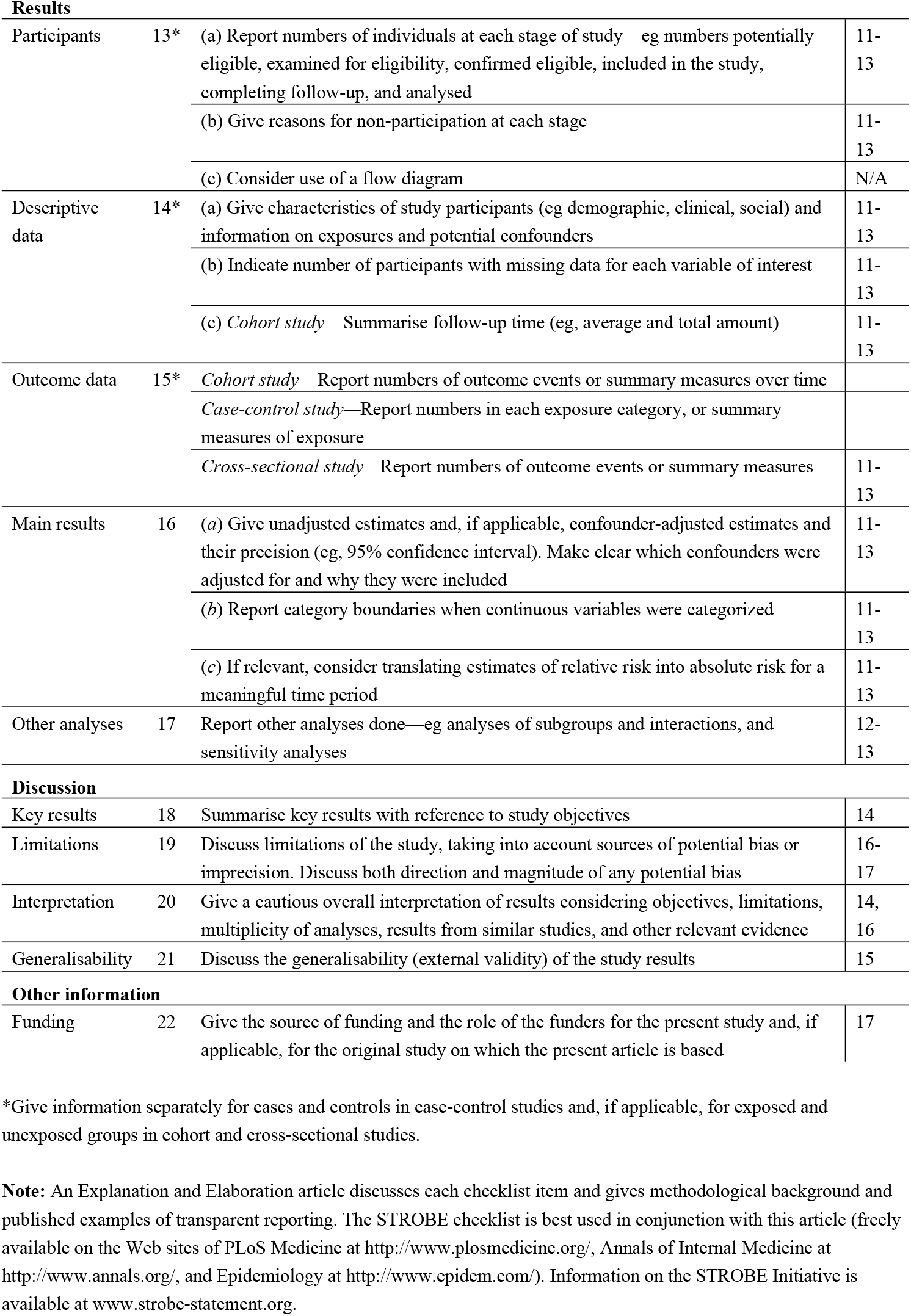

